# Leveraging Feature Transfer to Predict Medication Resistance and Secondary-Clinical Outcomes in Psychotic Disorders in Forensic Settings

**DOI:** 10.1101/2025.03.06.25321797

**Authors:** Devon Watts, Heather Moulden, Mini Mamak, Ives Passos, Gary Chaimowitz

**Affiliations:** Center for Precision Psychiatry, Massachusetts General Hospital, Boston, MA, USA; Department of Psychiatry and Behavioral Neurosciences, McMaster University, Hamilton, Canada; Laboratório de Molecular Psychiatry, Centro de Pesquisa Experimental (CPE) and Centro de Pesquisa Clínica (CPC), Hospital de Clínicas de Porto Alegre (HCPA), Porto Alegre, RS, Brasil

## Abstract

Medication resistance in psychotic disorders represents a critical challenge in forensic psychiatry, where up to 50% of patients show poor treatment response, leading to increased risk of relapse, violence, and rehospitalization. Feature Transfer, a novel machine learning framework based on rank aggregated feature selection, transfers predictive features identified for one outcome to related outcomes while maintaining clinical interpretability, a critical advantage over conventional transfer learning approaches that obscure feature level insights by transferring complex model parameters. Applied to psychotic disorders, this methodology identified key predictors for medication resistance and assessed their transferability to related clinical outcomes. Analyzing data from 893 patients across 11 forensic psychiatric institutions, we compared Feature Transfer models (using the top 25 features discriminating medication resistance from responders) with full feature models (95 features) for predicting clinical relapse, treatment non adherence, and escape behaviors. In the broader psychotic disorders sample, Feature Transfer achieved statistically equivalent performance to full feature models for clinical relapse and treatment non adherence (F1 score differences with confidence intervals overlapping zero), though performed less effectively for escape behaviors (AUC: 0.736 vs 0.838). In schizophrenia patients (n=634), Feature Transfer showed statistically significant improvement in F1 score for clinical relapse prediction compared to full feature models (difference: 0.119, 95% CI: 0.025 to 0.213), with notably higher sensitivity (0.912 vs 0.802) while maintaining comparable discriminative ability (AUC: 0.912 vs 0.925, difference not statistically significant). Treatment history features, particularly previous medication unresponsiveness and duration of clinical care, maintained high predictive importance across multiple clinical outcomes (relapse, non adherence, and escape behaviors), suggesting they represent fundamental risk indicators regardless of the specific outcome being predicted. While our retrospective design limits causal inference and relies on historical indicators as proxies for secondary outcomes (relapse and escape behaviors), the demonstrated utility of medication resistance features across different clinical outcomes reveals potential shared risk dimensions in psychotic disorders, particularly for relapse prediction in schizophrenia. Feature Transfer offers a transparent approach for identifying common predictive factors that could advance personalized intervention strategies in complex psychiatric populations.

## Introduction

Psychotic disorders, including schizophrenia (SCZ), bipolar disorder (BD), and schizoaffective disorder (SAD), are complex neuropsychiatric conditions characterized by a range of cognitive, emotional, and behavioral dysfunctions, including but not limited to, hallucinations, delusions, and disorganized thinking^1^. Each disorder presents unique challenges in both diagnosis and therapeutic intervention. Furthermore, medication resistance is a pervasive issue, posing significant barriers to effective symptom management in SCZ, BD, and SAD^2^, thereby complicating the clinical landscape across these conditions.

Considering the high prevalence of medication resistance in psychotic disorders, and complex symptomatology, forensic psychiatric institutions, which treat a high proportion of patients with severe psychotic symptoms, represents a critical setting for further investigation. For instance, in a recent meta-analysis, patients with SCZ in forensic settings showed higher symptom ratings across all positive and negative symptom severity (PANSS) subscales, including positive, negative, general, and total domains, relative to non-forensic patients^3^. Moreover, while mental disorders are more prevalent among incarcerated individuals than the general population^4^, schizophrenia spectrum disorders are particularly overrepresented in forensic psychiatric institutions worldwide^5^. To compound this issue, pseudo-resistance, a term that describes scenarios where treatment failure occurs due to factors such as treatment non-adherence, is a prevalent issue in SCZ, with rates estimated as high as 50%^5,6^. This not only impacts treatment efficacy but can also pose significant risks including clinical relapse and violence through poor symptom management^7,8^, increased rates of rehospitalization ^9^, and even suicide^9,10^. Therefore, there is a pressing need for novel methodologies that can disentangle these complexities. Conventional diagnostic and treatment paradigms often fall short in capturing the nuanced interplay of variables such as treatment resistance, non-adherence, and related clinical outcomes. Moreover, individual differences in outcomes across psychotic disorders remain difficult to predict based on clinical assessment alone^11^. Identifying which features are associated with multiple clinical outcomes could enable more efficient risk assessment and intervention planning, yet current approaches lack a systematic framework for evaluating shared predictive value across different clinical outcomes.

To address this, we propose a novel machine learning framework called Feature Transfer. This approach identifies the most predictive features for one task (in this case, medication resistance) and transfers their ranked importance to predict related tasks (such as clinical relapse, medication non-adherence, and escape behaviors), while maintaining interpretability. Feature Transfer draws inspiration from transfer learning and multi-task learning, but focuses on cross-task feature importance generalization rather than transferring model parameters or shared representations. Transfer learning typically adapts model parameters from a source task to improve performance on a target task^12^, while multi-task learning jointly optimizes shared representations across tasks^13^. However, these approaches may obscure the specific features driving predictions for individual outcomes. In contrast, Feature Transfer operates through rank-aggregated feature selection, where features identified as important for one task are empirically assessed for their predictive relevance across related outcomes. This aligns with emerging methodologies in transfer subspace learning, which seek to reduce domain discrepancy by preserving feature importance hierarchies^14^.

Feature Transfer offers a novel and systematic framework to evaluate the generalization of predictive features – encompassing not only traditional risk factors but also other clinically relevant markers – across related clinical outcomes in psychiatric prediction. This approach emphasizes feature importance in a way that has direct implications for clinical decision-making, helping clinicians understand and explain the basis for risk assessments and treatment decisions. When specific features maintain predictive relevance across multiple outcomes, they may represent key intervention targets that influence multiple aspects of illness trajectory. For example, if treatment history features strongly predict both medication resistance and clinical relapse, this suggests that early treatment experiences could be crucial intervention points. Similarly, if behavioral indicators consistently predict multiple clinical outcomes, this might guide the development of more extensive risk assessment protocols.

In contrast to traditional machine learning paradigms^15^, where separate predictive models are trained independently for each clinical outcome using either the complete feature set or outcome-specific feature selection, Feature Transfer allows for a more targeted method of knowledge transfer. Applied to psychotic disorders in this study, Feature Transfer addresses three main questions: (1) do features predictive of medication resistance generalize to other clinical outcomes; (2) does this generalization differ between broad psychotic disorders and SCZ specifically; and (3) can shared mechanisms or risk factors can be identified while maintaining model interpretability? The efficacy of this method depends on two key metrics: the similarity between tasks and the predictive power of transferred features. When transferred features achieve comparable or superior performance to full-feature models, this suggests shared risk factors or mechanisms between outcomes, offering valuable insights for tailoring clinical interventions.

In this study, we apply the Feature Transfer method to investigate the relationship between medication resistance and other clinically relevant outcomes in psychotic disorders. Our analyses were performed using a representative sample of 893 patients diagnosed with psychotic disorders (schizophrenia, bipolar disorder, and schizoaffective disorder) across 11 forensic psychiatric institutions in Ontario, Canada, including a subgroup analysis of 634 patients with schizophrenia. The methodological approach proceeds in two stages. First, we develop predictive models for medication resistance using random forest^16^ with recursive feature elimination (RFE)^17^ and an impurity-corrected importance measure^18^ to identify the most predictive features, while using super learner^19^ to assess their predictive performance through an ensemble of cross-validated algorithms. Second, we examine the transferability of these identified features to three additional clinical outcomes that frequently challenge forensic psychiatric care: clinical relapse, treatment non-adherence, and escape behaviors. These outcomes were selected based on their clinical significance and potential shared risk factors with medication resistance^20–21^, with feature transfer model performance and feature importance compared against full-feature models trained to predict each specific outcome.

## 2. Results

We applied the Feature Transfer framework to a cohort of 893 patients with psychotic disorders (including a schizophrenia-specific subgroup, *n* = 634) across 11 forensic psychiatric institutions in Ontario, Canada, to evaluate whether features predictive of medication resistance generalize to three clinically significant outcomes: clinical relapse, treatment non-adherence, and escape behaviors. The analytical pipeline integrated machine learning (random forest classifiers with recursive feature elimination) and ensemble methods (Super Learner) to compare full-feature (95 variables) and reduced-feature (25 variables) models (see Methods). Key performance metrics, including AUC, sensitivity, and specificity, demonstrated that the reduced-feature model retained predictive accuracy for medication resistance (AUC = 0.806 vs. 0.809 for full-feature models; Table 1), while Feature Transfer identified shared predictors across outcomes (Figures 2–5). Stratified bootstrap tests ^22^ (10,000 iterations) and McNemar’s analyses ^23^ confirmed robustness (Supplementary Tables S2–S4).

**Figure 1.**
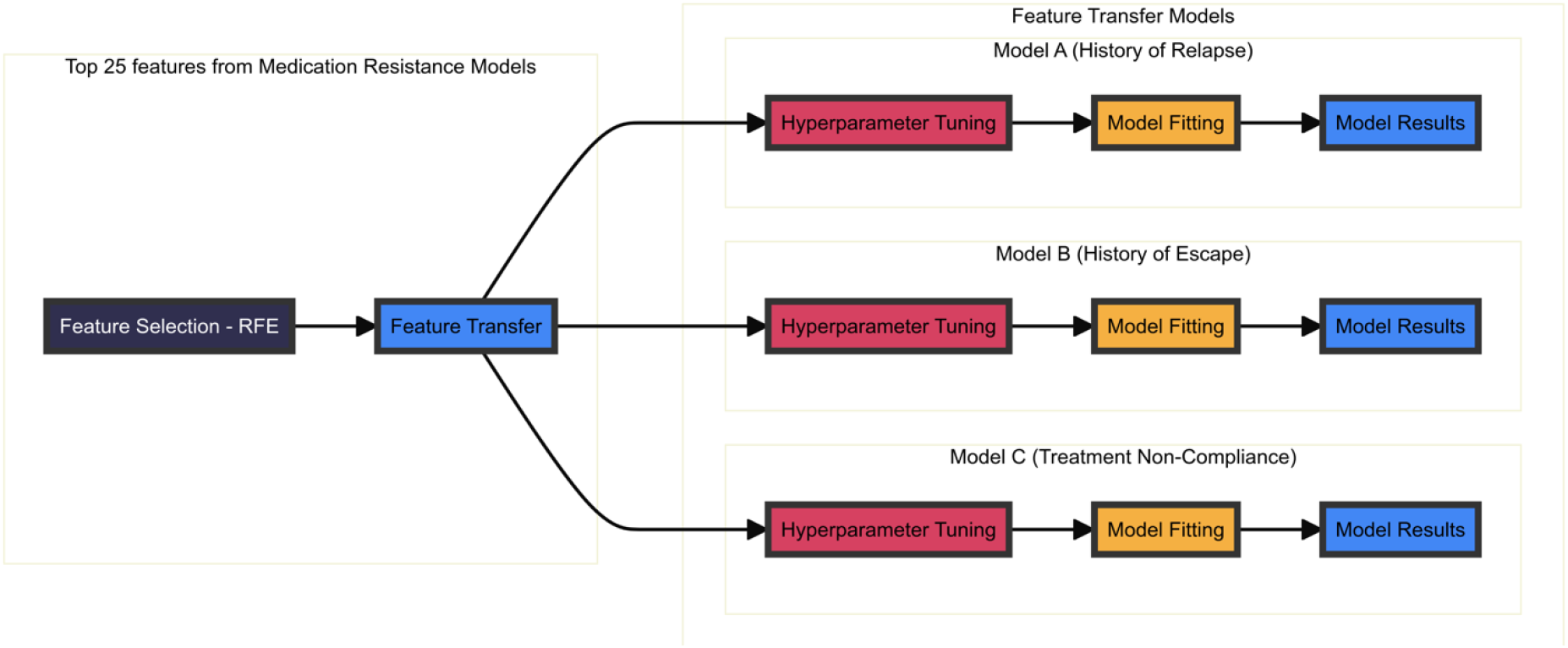
| Schematic of Feature Transfer Framework. The framework operates across three interconnected stages to investigate shared predictive factors between medication resistance and related clinical outcomes. Initially, Recursive Feature Elimination with a Random Forest classifier identifies the most predictive features for medication resistance, employing repeated cross-validation (5-fold, 5 repeats) to ensure robust feature selection. These identified features are then transferred to predict three distinct clinical outcomes: relapse history, escape behaviors, and medication non-compliance. Each prediction task utilizes Random Forest classifiers with Bayesian-optimized hyperparameters, enabling systematic evaluation of feature generalizability across outcomes. In parallel, Super Learner ensembles assess the maximum achievable predictive performance using these features, though at the cost of reduced interpretability. This dual modeling approach balances the need to identify interpretable shared risk factors while establishing benchmarks for predictive power across clinically-relevant outcomes in patients with psychotic disorders.

**Figure 2.**
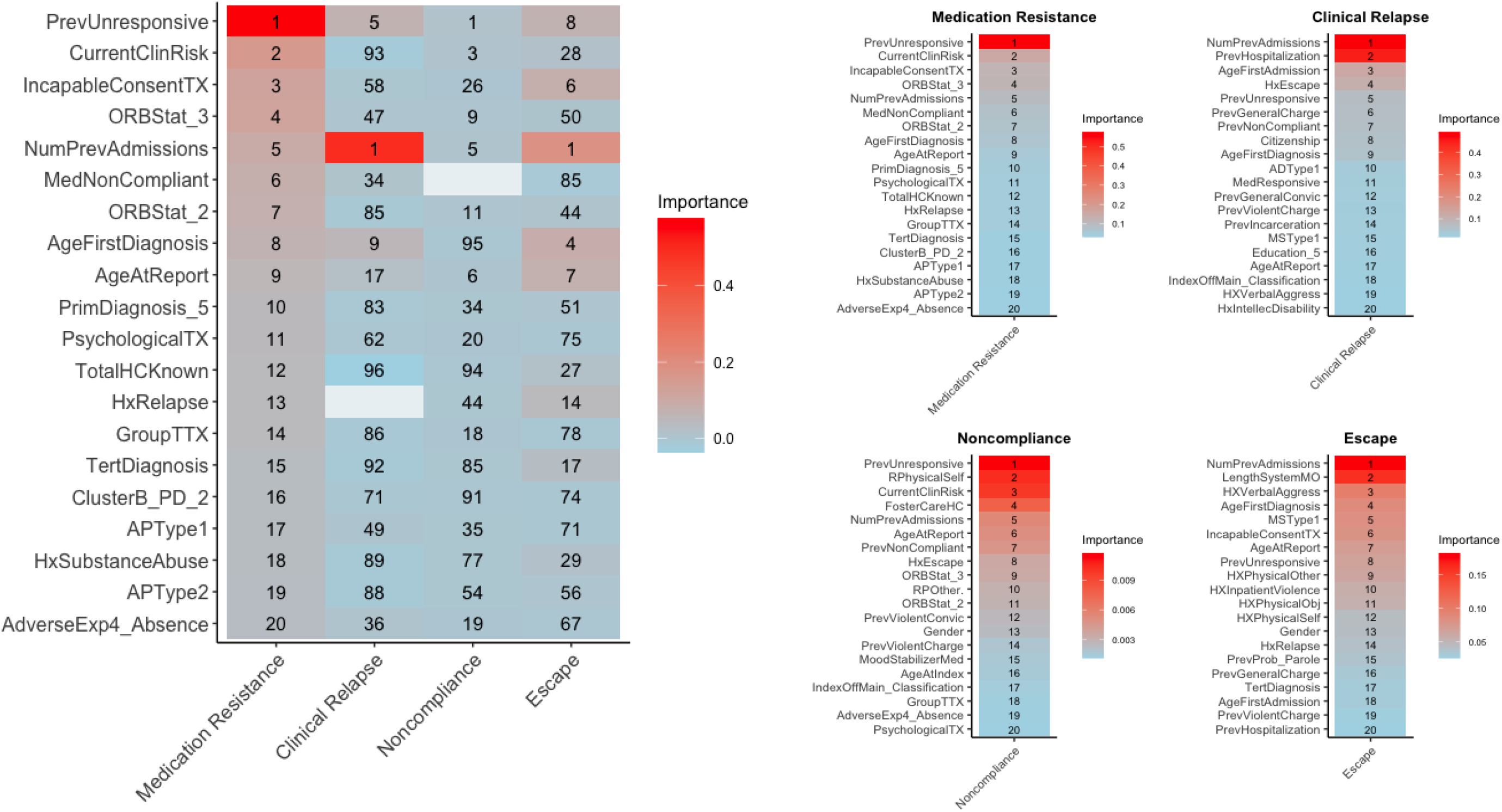
| Random forest variable importance ranking across data-driven models in psychotic disorders. This figure presents feature importance analysis from random forest classifiers using impurity-corrected importance measures across four clinical outcomes: medication resistance, clinical relapse, medication noncompliance, and escape behaviors. The *<u>left panel</u>* displays a heatmap of the top 20 features from the medication resistance model and their rankings across outcomes. APType1 and APType2 represent frequency-encoded primary and adjunctive antipsychotic prescription patterns. PrevUnresponsive indicates previous medication unresponsiveness, while CurrentClinRisk represents current clinical risk assessment. IncapableConsentTX denotes inability to consent to treatment. ORBStat reflects Ontario Review Board status, with NumPrevAdmissions tracking previous psychiatric admissions. AgeFirstDiagnosis and AgeAtReport capture timing of initial diagnosis and reporting. PrimDiagnosis and TertDiagnosis indicate primary and tertiary diagnostic classifications. PsychologicalTX denotes current psychological treatment status. White spaces in the heatmap indicate where features served as outcome variables. The color scale represents relative importance from 0.0 to 0.4, with darker red indicating higher importance. The *<u>right panel</u>* presents four separate heatmaps displaying the top 20 features by importance score for each outcome model. For medication resistance, treatment history features were the most influential features, with PrevUnresponsive ranking first, CurrentClinRisk second, and IncapableConsentTX third. The clinical relapse model identified institutional history as primary predictors, with NumPrevAdmissions and PrevHospitalization ranking first and second, followed by AgeFirstAdmission, HxEscape, and PrevUnresponsive. The noncompliance model prioritized previous treatment response patterns, with PrevUnresponsive ranking first, followed by HxPhysicalSelf and CurrentClinRisk. Legal status indicators and demographic factors appeared in middle rankings. The escape model emphasized institutional metrics, with NumPrevAdmissions and LengthSystemMO ranking first and second, followed by HXVerbalAggress. Treatment-related variables and various forms of aggressive behavior demonstrated strong predictive value in middle rankings. Importance score scales varied between outcomes, ranging from 0.1-0.4 for medication resistance to 0.05-0.15 for escape behaviors, reflecting outcome-specific distributions in feature importance. Feature rankings showed notable patterns across outcomes. PrevUnresponsive ranked first for both medication resistance and noncompliance but eighth for escape. CurrentClinRisk demonstrated substantial variability, ranking second for medication resistance but 93rd for clinical relapse. NumPrevAdmissions maintained consistent high importance, ranking fifth for medication resistance and first for clinical relapse, demonstrating its broad predictive relevance across multiple outcomes.

**Table 1.** | Performance of Medication Resistance Models in Psychotic Disorders. A comparison of performance metrics between a random forest model trained using the total A comparison of performance metrics between two random forest classifiers in discriminating medication resistance from medication responders among individuals with psychotic disorders. The dataset comprised 893 patients from 11 psychiatric institutions, with 39.8% classified as medication-resistant (*n*=355) and 60.2% as medication-responsive (*n*=538). This distribution was maintained through stratified sampling in both training (*n*=624; 248 resistant, 376 responsive) and testing (*n*=269; 107 resistant, 162 responsive) sets. Two models were developed and compared: 1) a model utilizing all 95 available features after preprocessing, 2) a reduced model utilizing only the top 25 features identified through recursive feature elimination (RFE) with a random forest model in the training set, with 5-fold cross-validation and 5-repeats. Both models underwent Bayesian optimization for hyperparameter tuning, a technique that efficiently balances exploitation of known high-performing regions and exploration of unexplored parameter spaces. As shown in Table 1, the results demonstrate comparable predictive performance between the two models across several key metrics. The model using all 95 features achieved an F1 score of 0.745, while the RFE model with 25 features attained an F1 score of 0.775. Sensitivity was 0.697 (95% CI: 0.619-0.765) in the full model and 0.747 (95% CI: 0.671-0.810) in the RFE model. Specificity showed comparable performance, with 0.738 (95% CI: 0.643-0.816) for the full model and 0.729 (95% CI: 0.633-0.808) for the RFE model. The positive predictive value (PPV) was 0.801 (95% CI: 0.724-0.862) in the full model and 0.806 (95% CI: 0.732-0.864) in the RFE model, while the negative predictive value (NPV) was 0.617 (95% CI: 0.527-0.700) in the full model and 0.655 (95% CI: 0.562-0.738) in the RFE model. Both models demonstrated similar discriminative ability, with the full model achieving an AUC of 0.809 (95% CI: 0.758-0.859) and the RFE model reaching 0.806 (95% CI: 0.755-0.857). The overlapping confidence intervals across all metrics suggest comparable performance between the two models, with the RFE model showing a trend towards slightly better performance in most areas.

| Medication Resistance | Psychotic Disorders<br>(All features – 95) | Psychotic Disorders<br>(Top 25 Features RFE) |
| --- | --- | --- |
| F1 Score | 0.745 | 0.775 |
| Sensitivity | 0.697 (95% CI: 0.619-0.765) | 0.747 (95% CI: 0.671-0.810) |
| Specificity | 0.738 (95% CI: 0.643-0.816) | 0.729 (95% CI: 0.633-0.808) |
| PPV | 0.801 (95% CI: 0.724-0.862) | 0.806 (95% CI: 0.732-0.864) |
| NPV | 0.617 (95% CI: 0.527-0.700) | 0.655 (95% CI: 0.562-0.738) |
| AUC | 0.809 (95% CI: 0.758-0.859) | 0.806 (95% CI: 0.755-0.857) |
| PRAUC | 0.876 (95% CI: 0.833-0.912) | 0.874 (95% CI: 0.832-0.910) |

### 2.1. Reduced feature sets maintain predictive accuracy for medication resistance

First, we compared Random forest models using the full set of 95 features to those using the top 25 RFE-selected features for predicting medication resistance in patients with psychotic disorders (*n* = 893) and a subgroup of individuals with schizophrenia (*n* = 634). In the broader psychotic disorders sample (**Table 1**), the reduced-feature model achieved similar discriminative ability to the full-feature model (AUC: 0.806, 95% CI: 0.755-0.857 vs 0.809, 95% CI: 0.758-0.859). Minor differences emerged across other metrics, with the reduced-feature model showing a slightly higher F1 score (0.775 vs 0.745) and sensitivity (0.747 [95% CI: 0.671-0.810] vs 0.697 [95% CI: 0.619-0.765]), correctly identifying about 75 versus 70 of every 100 truly medication-resistant patients. Specificity was similarly maintained (0.729 [95% CI: 0.633-0.808] vs 0.738 [95% CI: 0.643-0.816]), consistently identifying approximately 73 of every 100 truly medication-responsive patients, and positive predictive values remained around 80% (0.806 [95% CI: 0.732-0.864] vs 0.801 [95% CI: 0.724-0.862]). Negative predictive values differed marginally (0.617 [95% CI: 0.527-0.700] vs 0.655 [95% CI: 0.562-0.738]). Although overlapping confidence intervals preclude definitive conclusions, these findings suggest that retaining approximately one-quarter of the original features is sufficient to maintain predictive accuracy for medication resistance in psychotic disorders.

In the schizophrenia subgroup (**Table 2**), the reduced-feature model demonstrated higher point estimates across several metrics relative to the full-feature model. For example, AUC increased from 0.735 (95% CI: 0.666–0.804) to 0.824 (95% CI: 0.767–0.882), and sensitivity rose from 0.687 (95% CI: 0.593–0.768) to 0.756 (95% CI: 0.667–0.829), correctly identifying about 76 versus 69 of every 100 truly medication-resistant patients. Specificity improved from 0.592 (95% CI: 0.473–0.702) to 0.671 (95% CI: 0.553–0.772), and positive predictive value increased from 0.741 (95% CI: 0.647–0.819) to 0.784 (95% CI: 0.695–0.854). The F1 score rose from 0.702 to 0.766, suggesting that the reduced set may enhance predictive performance for medication resistance in schizophrenia. However, these improvements should be interpreted cautiously given overlapping confidence intervals. Subsequent analyses employing stratified bootstrap tests and McNemar’s tests further examined whether features identified for medication resistance prediction could effectively generalize to secondary outcomes (clinical relapse, treatment non-adherence, and escape behaviors).

**Table 2.** | Performance of Medication Resistance Classifiers in Schizophrenia. A comparison of performance metrics between two random forest classifiers in discriminating medication resistance from medication responders among individuals with Schizophrenia. This model comprised 634 individuals with schizophrenia from 11 psychiatric institutions, with 443 individuals allocated to the training set and 191 to the testing set. Stratified sampling was employed to maintain proportional representation of medication-resistant and medication-responsive individuals in both the training and testing sets, ensuring that each subset accurately reflected the overall population distribution. The dataset comprised 634 individuals with schizophrenia, with 39.8% classified as medication-resistant (*n* = 252) and 60.2% as medication-responsive (*n* = 382). This distribution was maintained through stratified sampling in both training (443 individuals; 39.7% resistant) and testing (191 individuals; 39.8% resistant) sets. Two models were developed and compared, comprising a model with all 95 features available after preprocessing, and a reduced model using the top 25 features identified through recursive feature elimination (RFE). The models were trained on a sample of 443 individuals with psychotic disorders, across 11 psychiatric institutions, and evaluated on a testing set of 191 individuals, using stratified sampling to ensure balanced class representation. Hyperparameters were optimized using Bayesian optimization to enhance model performance. As shown in Table 2, the performance metrics reveal differences between the full feature model and the Recursive Feature Elimination (RFE) model, although overlapping confidence intervals suggest these differences may not be statistically significant. Notably, the RFE model, using only 26.3 percent of the original features, demonstrates comparable performance to the full model, which has implications for model interpretability and efficiency. The RFE model consistently shows a trend towards better performance across all metrics. Both models perform better than chance, with the RFE model indicating a trend towards superior overall discriminative ability. However, the overlapping confidence intervals necessitate cautious interpretation of these differences.

| Performance Metrics | Medication Resistance – Schizophrenia (All features – 95) | Medication Resistance – Schizophrenia (Top 25 Features RFE) |
| --- | --- | --- |
| F1 Score | 0.702 | 0.766 |
| Sensitivity | 0.687 (95% CI: 0.593-0.768) | 0.756 (95% CI: 0.667, 0.829) |
| Specificity | 0.592 (95% CI: 0.473-0.702) | 0.671 (95% CI: 0.553, 0.772) |
| PPV | 0.718 (95% CI: 0.623-0.798) | 0.776 (95% CI: 0.686, 0.848) |
| NPV | 0.555 (95% CI: 0.441-0.664) | 0.645 (95% CI: 0.529, 0.747) |
| AUC | 0.735 (95% CI: 0.666-0.804) | 0.824 (95% CI: 0.767, 0.882) |
| PRAUC | 0.826 (95% CI: 0.763-0.878) | 0.876 (95% CI: 0.819, 0.921) |

### 2.2. Medication resistance features successfully transfer to predict clinical relapse

To assess the extent to which features predictive of medication resistance generalized to other clinical outcomes, we compared Feature Transfer Models (using the top 25 medication resistance features) with Outcome-Specific Models (using all 95 features) for predicting clinical relapse, medication non-compliance, and escape behaviors. These comparisons were conducted in the full psychotic disorders sample (*n* = 893) and in a schizophrenia-specific subgroup (*n* = 634). In the psychotic disorders sample (**Table 3**), both model types produced high sensitivity and positive predictive value when predicting History of Clinical Relapse (e.g., Feature Transfer Model sensitivity: 0.879 [95% CI: 0.828-0.907]; Outcome-Specific Model sensitivity: 0.866 [95% CI: 0.814-0.906]). While the Outcome-Specific Model had a slightly higher AUC point estimate (0.889 vs 0.862), the bootstrap-derived difference in AUC included zero (difference: -0.011 [95% CI: -0.112, 0.081]) and was therefore not statistically significant. Similarly, the difference in F1 scores also included zero (difference: -0.007 [95% CI: -0.083, 0.069]). In contrast, for History of Escape, the differences in both AUC and F1 score excluded zero (AUC difference: -0.160 [95% CI: -0.304, -0.031]; F1 difference: -0.706 [95% CI: -0.742, -0.668]), indicating that the Outcome-Specific Model had statistically significantly higher performance. For Medication Noncompliance, both F1 and AUC differences included zero, suggesting no statistically significant difference between models. McNemar’s tests further supported these findings: there were significant classification decision differences for History of Relapse (χ² = 12.04, p = 0.0005) and History of Escape (χ² = 16.00, p < 0.0001), but not for Medication Noncompliance (χ² = 1.33, p = 0.248).

**Table 3.**
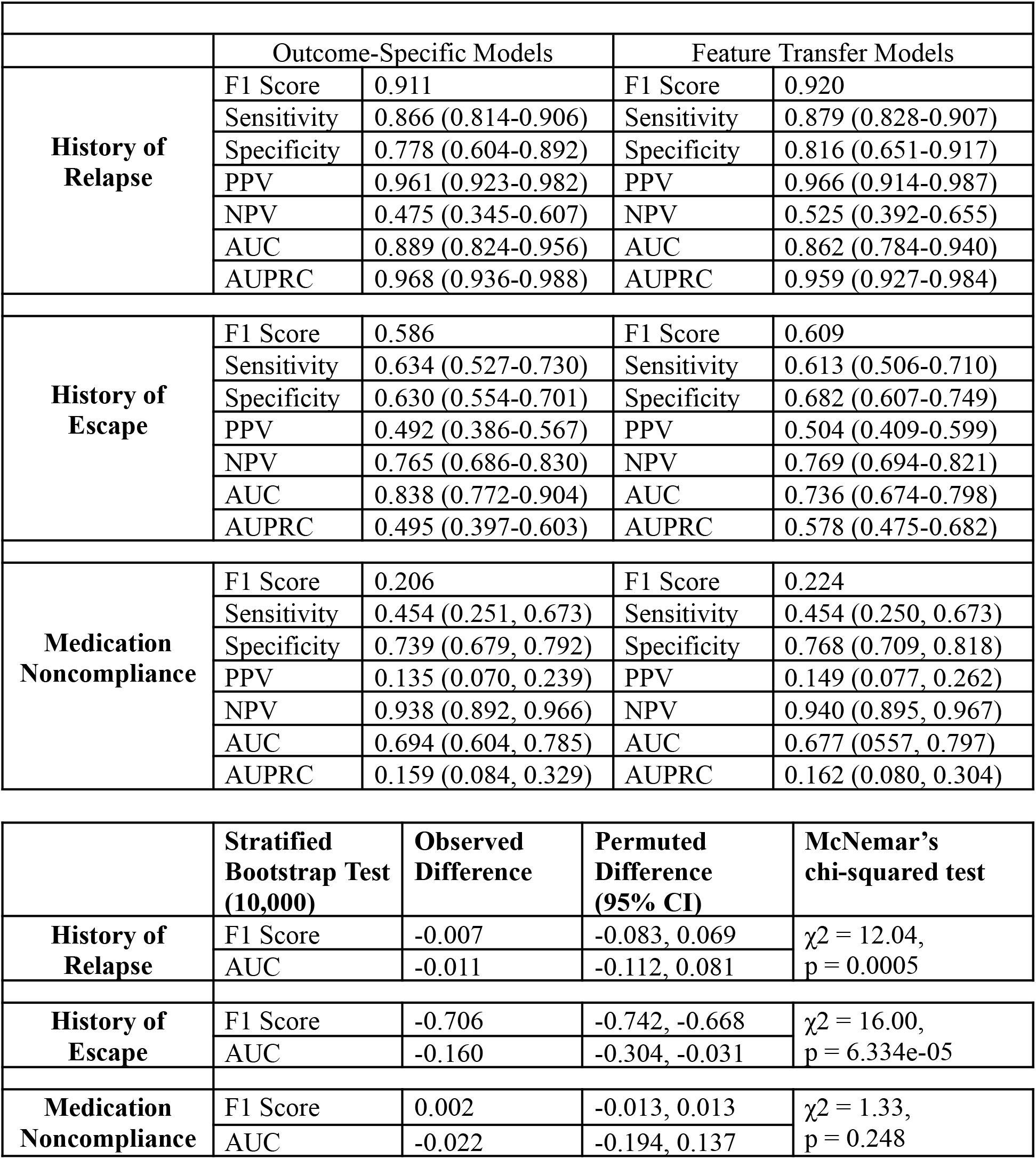
| Performance of Outcome-Specific and Feature Transfer Models in Psychotic Disorders. This table presents performance metrics comparing Outcome-Specific Models (95 features) and Feature Transfer Models (25 medication resistance-derived features) across three clinical outcomes in patients with psychotic disorders (*n* = 893). The dataset distributions were: History of Relapse (positive class: 86.6%, n=773), History of Escape (positive class: 34.5%, n=308), and Medication Noncompliance (positive class: 8.8%, n=79). These distributions were maintained through stratified sampling in training (*n* = 624) and testing (*n* = 269) sets. For Medication Noncompliance, majority class downsampling (ratio=0.3) was applied during training while preserving the original distribution in the test set. For History of Relapse, both models achieved comparable metrics. The Feature Transfer Model showed an F1 Score of 0.920 and AUC of 0.862 (95% CI: 0.784-0.940), while the Outcome-Specific Model achieved an F1 Score of 0.911 and AUC of 0.889 (95% CI: 0.824-0.956). For History of Escape, the Feature Transfer Model’s F1 Score was 0.609 compared to the Outcome-Specific Model’s 0.586, though the Outcome-Specific Model demonstrated higher AUC (0.838, 95% CI: 0.772-0.904 vs 0.736, 95% CI: 0.674-0.798). For Medication Noncompliance, both models showed similar performance metrics, with Feature Transfer Model F1 Score at 0.224 versus 0.206 for the Outcome-Specific Model, and comparable AUC values (0.677, 95% CI: 0.557-0.797 vs 0.694, 95% CI: 0.604-0.785). Statistical comparison using stratified bootstrap tests (10,000 resamples) indicated minimal differences for History of Relapse (F1 Score difference: -0.007, 95% CI: -0.083 to 0.069; AUC difference: -0.011, 95% CI: -0.112 to 0.081) and Medication Noncompliance (F1 Score difference: 0.002, 95% CI: -0.013 to 0.013; AUC difference: -0.022, 95% CI: -0.194 to 0.137). History of Escape showed larger differences (F1 Score difference: -0.706, 95% CI: -0.742 to -0.668; AUC difference: -0.160, 95% CI: -0.304 to -0.031). McNemar’s chi-squared test revealed significant classification differences for History of Relapse (χ2 = 12.04, p = 0.0005) and History of Escape (χ2 = 16.00, p = 6.334e-05), but not for Medication Noncompliance (χ2 = 1.33, p = 0.248), indicating distinct classification patterns between model types despite similar aggregate performance metrics.

|  | Outcome-Specific Models |  | Feature Transfer Models |  |
| --- | --- | --- | --- | --- |
| History of Relapse | F1 Score | 0.911 | F1 Score | 0.920 |
|  | Sensitivity | 0.866 (0.814-0.906) | Sensitivity | 0.879 (0.828-0.907) |
|  | Specificity | 0.778 (0.604-0.892) | Specificity | 0.816 (0.651-0.917) |
|  | PPV | 0.961 (0.923-0.982) | PPV | 0.966 (0.914-0.987) |
|  | NPV | 0.475 (0.345-0.607) | NPV | 0.525 (0.392-0.655) |
|  | AUC | 0.889 (0.824-0.956) | AUC | 0.862 (0.784-0.940) |
|  | AUPRC | 0.968 (0.936-0.988) | AUPRC | 0.959 (0.927-0.984) |
| History of Escape | F1 Score | 0.586 | F1 Score | 0.609 |
|  | Sensitivity | 0.634 (0.527-0.730) | Sensitivity | 0.613 (0.506-0.710) |
|  | Specificity | 0.630 (0.554-0.701) | Specificity | 0.682 (0.607-0.749) |
|  | PPV | 0.492 (0.386-0.567) | PPV | 0.504 (0.409-0.599) |
|  | NPV | 0.765 (0.686-0.830) | NPV | 0.769 (0.694-0.821) |
|  | AUC | 0.838 (0.772-0.904) | AUC | 0.736 (0.674-0.798) |
|  | AUPRC | 0.495 (0.397-0.603) | AUPRC | 0.578 (0.475-0.682) |
| Medication Noncompliance | F1 Score | 0.206 | F1 Score | 0.224 |
|  | Sensitivity | 0.454 (0.251, 0.673) | Sensitivity | 0.454 (0.250, 0.673) |
|  | Specificity | 0.739 (0.679, 0.792) | Specificity | 0.768 (0.709, 0.818) |
|  | PPV | 0.135 (0.070, 0.239) | PPV | 0.149 (0.077, 0.262) |
|  | NPV | 0.938 (0.892, 0.966) | NPV | 0.940 (0.895, 0.967) |
|  | AUC | 0.694 (0.604, 0.785) | AUC | 0.677 (0.557, 0.797) |
|  | AUPRC | 0.159 (0.084, 0.329) | AUPRC | 0.162 (0.080, 0.304) |

**Table 3 | Performance of Outcome-Specific and Feature Transfer Models in Psychotic Disorders.**
|  | Stratified Bootstrap Test (10,000) | Observed Difference | Permuted Difference (95% CI) | McNemar's chi-squared test |
| --- | --- | --- | --- | --- |
| History of Relapse | F1 Score | -0.007 | -0.083, 0.069 | $\chi^2 = 12.04$,<br>$p = 0.0005$ |
|  | AUC | -0.011 | -0.112, 0.081 |  |
| History of Escape | F1 Score | -0.706 | -0.742, -0.668 | $\chi^2 = 16.00$,<br>$p = 6.334e-05$ |
|  | AUC | -0.160 | -0.304, -0.031 |  |
| Medication Noncompliance | F1 Score | 0.002 | -0.013, 0.013 | $\chi^2 = 1.33$,<br>$p = 0.248$ |
|  | AUC | -0.022 | -0.194, 0.137 |  |

In the schizophrenia subgroup (**Table 4**), stratified bootstrap results indicated a statistically significant improvement in F1 score for History of Relapse when using the Feature Transfer Model (difference: 0.119, 95% CI: 0.025 to 0.213), while the Outcome-Specific Model showed a significant advantage for Medication Noncompliance (difference: -0.299, 95% CI: -0.330 to -0.271). No significant F1 score difference emerged for History of Escape (difference: -0.011, 95% CI: -0.083 to 0.108). AUC differences for all outcomes included zero, suggesting that discriminative ability was broadly similar at the aggregate level. McNemar’s tests revealed that classification decisions differed significantly for History of Relapse (χ² = 23.18, p < 0.0001) and Medication Noncompliance (χ² = 9.521, p = 0.002), but not for History of Escape (χ² = 0.923, p = 0.337). Notably, the Feature Transfer model’s ability to match or improve upon the Outcome-Specific model performance with fewer features suggests that a subset of core predictors, originally identified for medication resistance, can effectively capture key risk dimensions across multiple clinical outcomes in schizophrenia—especially for predicting relapse, where the Feature Transfer model achieved higher sensitivity (0.912 vs 0.802) without a statistically significant reduction in AUC.

**Table 4.** | Performance of Outcome-Specific and Feature Transfer Models in Schizophrenia. This table presents a comparative analysis of predictive performance between Outcome-Specific Models and Feature Transfer Models for three clinically relevant outcomes in patients with schizophrenia (*n* = 634). The dataset’s class distributions were: History of Relapse (positive class: 85.2%, n=540), History of Escape (positive class: 33.4%, n=212), and Medication Noncompliance (positive class: 8.2%, n=52). These proportions were preserved through stratified sampling in both training (*n* = 443) and testing (*n* = 191) sets. For Medication Noncompliance, we applied majority class downsampling (ratio=0.3) during model training while maintaining the original distribution in the test set. The Outcome-Specific Models were trained using all 95 available features, while the Feature Transfer Models utilized the top 25 features identified as most predictive of medication resistance in schizophrenia. For History of Relapse, the Feature Transfer Model demonstrated slightly superior performance compared to the Outcome-Specific Model. The Feature Transfer Model achieved a higher F1 Score (0.932 vs. 0.878) and comparable AUC (0.912, 95% CI: 0.850-0.974 vs. 0.925, 95% CI: 0.871-0.979). Notably, the Feature Transfer Model showed higher sensitivity (0.912 vs. 0.802) with overlapping specificity (0.827 vs. 0.862). The stratified bootstrap test revealed a significant difference in F1 Scores (observed difference: 0.119, 95% CI: 0.025-0.213), while the difference in AUC was not significant (observed difference: -0.013, 95% CI: -0.095-0.066). The McNemar’s test showed a significant difference in classifications (χ2 = 23.18, p = 1.469e-06), suggesting that the Feature Transfer Model captures different aspects of relapse risk compared to the Outcome-Specific Model. For History of Escape, both models showed moderate performance with similar F1 Scores (Feature Transfer: 0.566, Outcome-Specific: 0.525) and AUCs (Feature Transfer: 0.664, 95% CI: 0.584-0.774; Outcome-Specific: 0.684, 95% CI: 0.607-0.761). The Feature Transfer Model demonstrated higher sensitivity (0.734 vs. 0.565) but lower specificity (0.567 vs. 0.708). The stratified bootstrap test showed no significant differences in F1 Scores (observed difference: -0.011, 95% CI: -0.083-0.108) or AUCs (observed difference: -0.020, 95% CI: -0.131-0.093). The McNemar’s test was not significant (χ2 = 0.923, p = 0.3367), indicating similar classification patterns between the models. For Medication Noncompliance, both models exhibited relatively low performance. The Feature Transfer Model showed a slightly higher F1 Score (0.209 vs. 0.174) and AUC (0.631, 95% CI: 0.477-0.785 vs. 0.598, 95% CI: 0.457-0.739). The Feature Transfer Model demonstrated lower sensitivity (0.466 vs. 0.600) but higher specificity (0.744 vs. 0.551). The stratified bootstrap test revealed a significant difference in F1 Scores (observed difference: -0.299, 95% CI: -0.330--0.271), while the difference in AUCs was not significant (observed difference: -0.008, 95% CI: -0.215-0.156). McNemar’s test was significant (χ2 = 9.521, p = 0.0020), suggesting differences in classification patterns between the models. These results highlight the potential of the Feature Transfer approach in leveraging medication resistance predictors for related outcomes in schizophrenia. The comparable or slightly superior performance of Feature Transfer Models, particularly for relapse prediction, suggests shared risk factors between medication resistance and other clinical outcomes in schizophrenia. However, the variability in performance across outcomes underscores the complex nature of schizophrenia and the potential need for outcome-specific predictive models in certain cases.

|  | Outcome-Specific Models |  |  | Feature Transfer Models |  |
| --- | --- | --- | --- | --- | --- |
| History of Relapse | F1 Score | 0.878 |  | F1 Score | 0.932 |
|  | Sensitivity | 0.802 (0.731, 0.859) |  | Sensitivity | 0.912 (0.842, 0.941) |
|  | Specificity | 0.862 (0.674, 0.955) |  | Specificity | 0.827 (0.635, 0.935) |
|  | PPV | 0.970 (0.921, 0.990) |  | PPV | 0.969 (0.920, 0.987) |
|  | NPV | 0.438 (0.309, 0.576) |  | NPV | 0.600 (0.434, 0.747) |
|  | AUC | 0.925 (0.871, 0.979) |  | AUC | 0.912 (0.850, 0.974) |
|  | AUPRC | 0.977 (0.960, 0.999) |  | AUPRC | 0.836 (0.756, 0.900) |
| History of Escape | F1 Score | 0.525 |  | F1 Score | 0.566 |
|  | Sensitivity | 0.565 (0.433, 0.684) |  | Sensitivity | 0.734 (0.607, 0.833) |
|  | Specificity | 0.708 (0.620, 0.784) |  | Specificity | 0.567 (0.456, 0.654) |
|  | PPV | 0.493 (0.375, 0.612) |  | PPV | 0.461 (0.363, 0.562) |
|  | NPV | 0.763 (0.674, 0.834) |  | NPV | 0.809 (0.709, 0.882) |
|  | AUC | 0.684 (0.607, 0.761) |  | AUC | 0.664 (0.584, 0.774) |
|  | AUPRC | 0.463 (0.358, 0.597) |  | AUPRC | 0.314 (0.210, 0.367) |
| Medication Noncompliance | F1 Score | 0.174 |  | F1 Score | 0.209 |
|  | Sensitivity | 0.600 (0.329, 0.825) |  | Sensitivity | 0.466 (0.223, 0.726) |
|  | Specificity | 0.551 (0.474, 0.625) |  | Specificity | 0.744 (0.672, 0.805) |
|  | PPV | 0.102 (0.050, 0.189) |  | PPV | 0.135 (0.060, 0.264) |
|  | NPV | 0.941 (0.872, 0.976) |  | NPV | 0.942 (0.886, 0.973) |
|  | AUC | 0.598 (0.457, 0.739) |  | AUC | 0.631 (0.477, 0.785) |
|  | AUPRC | 0.073 (0.039, 0.159) |  | AUPRC | 0.107 (0.051, 0.238) |

**Table 4 | Performance of Outcome-Specific and Feature Transfer Models in Schizophrenia.**
|  | Stratified Bootstrap Test (10,000) | Observed Difference | Permuted Difference (95% CI) | McNemar's chi-squared test |
| --- | --- | --- | --- | --- |
| History of Relapse | F1 Score | 0.119 | 0.025, 0.213 | $\chi^2 = 23.18$,<br>$p = 1.469\text{e-}06$ |
|  | AUC | -0.013 | -0.095, 0.066 |  |
| History of Escape | F1 Score | -0.011 | -0.083, 0.108 | $\chi^2 = 0.923$,<br>$p = 0.3367$ |
|  | AUC | -0.020 | -0.131, 0.093 |  |
| Medication Noncompliance | F1 Score | -0.299 | -0.330, -0.271 | $\chi^2 = 9.521$,<br>$p = 0.0020$ |
|  | AUC | -0.008 | -0.215, 0.156 |  |

### 2.3. Super learner and random forest models show variable performance across outcomes

To benchmark the maximum achievable predictive performance while maintaining feature interpretability, we compared a super learner framework to our random forest classifiers, where both were trained using the top 25 features identified for medication resistance (**Tables 3-5**). In the psychotic disorder sample, the super learner model demonstrated lower F1 score (0.650) compared to the random forest Feature Transfer model (0.920), with comparable AUC values (super learner: 0.815 [95% CI: 0.762-0.861] vs random forest: 0.862 [95% CI: 0.784-0.940]). For secondary outcomes, performance patterns varied. For History of Relapse in psychotic disorders, both approaches achieved strong performance (super learner F1: 0.954, AUC: 0.881 [95% CI: 0.793-0.952]; random forest F1: 0.920, AUC: 0.862 [95% CI: 0.784-0.940]). However, for History of Escape, the super learner showed lower performance (F1: 0.455, AUC: 0.676 [95% CI: 0.607-0.742]) compared to the random forest model (F1: 0.609, AUC: 0.736 [95% CI: 0.674-0.798]). In the schizophrenia sample, while both approaches showed comparable AUC values for medication resistance prediction (super learner: 0.789 [95% CI: 0.724-0.848]; random forest: 0.912 [95% CI: 0.850-0.974]), performance varied for secondary outcomes. For Medication Noncompliance, despite the super learner achieving a higher F1 score (0.838 vs 0.209), the AUC values were similar (super learner: 0.583 [95% CI: 0.467-0.723]; random forest: 0.631 [95% CI: 0.477-0.785]) with overlapping confidence intervals, suggesting no clear advantage of either approach.

**Table 5.** | Performance of super learner classifiers using top 25 features of medication resistance across outcomes. This table presents the performance metrics of Super Learner models applied to medication resistance prediction and related clinical outcomes in two cohorts. The psychotic disorders cohort (*n* = 893) comprised the following class distributions: History of Relapse (86.6% positive, n=773), History of Escape (34.5% positive, n=308), and Medication Noncompliance (8.8% positive, n=79). The schizophrenia cohort (*n* = 634) showed similar distributions: History of Relapse (85.2% positive, n=540), History of Escape (33.4% positive, n=212), and Medication Noncompliance (8.2% positive, n=52). These proportions were maintained through stratified sampling in training and testing sets for both cohorts. Given the notable class imbalance, Medication Noncompliance models in both cohorts employed majority class downsampling (ratio=0.3) during training while preserving original distributions in test sets. The Super Learner ensemble utilized the top 25 features identified as most predictive of medication resistance through Recursive Feature Elimination. The Super Learner framework incorporated six diverse base learners: Random Forest, Bayesian Adaptive Regression Trees (BART), Support Vector Machine (SVM), Elastic Net, k-Nearest Neighbors (kNN), and Naive Bayes. These base learners were chosen to capture a range of underlying data patterns and relationships. An XGBoost model served as the meta-learner, leveraging the predictions from the base learners to generate final predictions. For the psychotic disorder sample, the Super Learner demonstrated strong performance in predicting History of Relapse (F1 Score: 0.954, 95% CI: 0.934-0.972; AUC: 0.881, 95% CI: 0.793-0.952). However, its performance was more modest for Medication Resistance prediction (F1 Score: 0.650, 95% CI: 0.570-0.722; AUC: 0.815, 95% CI: 0.762-0.861) and notably lower for History of Escape and Medication Noncompliance. In the schizophrenia sample, the Super Learner showed comparable overall performance for Medication Resistance prediction (F1 Score: 0.663, 95% CI: 0.575-0.738; AUC: 0.789, 95% CI: 0.724-0.848) relative to the psychotic disorders cohort. Notably, its improved F1 Score for Medication Noncompliance (0.838 vs. 0.298) was related to improvements in sensitivity (0.727 vs. 0.357) and PPV (0.925 vs. 0.256), at the expense of specificity (0.226 vs. 0.879) and NPV (0.089 vs. 0.921), indicating that while the model became more adept at identifying noncompliant patients, it simultaneously increased false positives. A similar pattern emerged for History of Escape: the rise in F1 Score (0.753 vs. 0.455) was driven by improved sensitivity (0.819 vs. 0.376) and PPV (0.698 vs. 0.574) but accompanied by a notable decline in specificity (0.297 vs. 0.852) and NPV (0.452 vs. 0.721). Taken together, these findings suggest that the clinical context of schizophrenia may enable certain medication resistance features to more effectively flag positive cases—even for challenging outcomes like noncompliance—though it does so at the cost of distinguishing them from negative cases.

| Super Learner Classifiers – Psychotic Disorders (N = 893) |  |  |  |  |
| --- | --- | --- | --- | --- |
|  | Medication Resistance | History of Relapse | History of Escape | Medication Noncompliance |
| F1 Score | 0.650 (0.570, 0.722) | 0.954 (0.934, 0.972) | 0.455 (0.354, 0.548) | 0.298 (0.148, 0.438) |
| Sensitivity | 0.626 (0.533, 0.718) | 0.958 (0.920, 0.975) | 0.376 (0.446, 0.698) | 0.357 (0.122, 0.400) |
| Specificity | 0.802 (0.739, 0.862) | 0.600 (0.458, 0.818) | 0.852 (0.660, 0.780) | 0.879 (0.885, 0.955) |
| PPV | 0.677 (0.582, 0.767) | 0.950 (0.932, 0.983) | 0.574 (0.279, 0.474) | 0.256 (0.182, 0.542) |
| NPV | 0.764 (0.699, 0.828) | 0.643 (0.419, 0.773) | 0.721 (0.797, 0.902) | 0.921 (0.836, 0.918) |
| AUC | 0.815 (0.762, 0.861) | 0.881 (0.793, 0.952) | 0.676 (0.607, 0.742) | 0.673 (0.560, 0.773) |
| AUPRC | 0.691 (0.596, 0.783) | 0.975 (0.948, 0.994) | 0.511 (0.411, 0.621) | 0.198 (0.111, 0.347) |
| Super Learner Classifiers – Schizophrenia (N = 634) |  |  |  |  |
|  | Medication Resistance | History of Relapse | History of Escape | Medication Noncompliance |
| F1 Score | 0.663 (0.575, 0.738) | 0.687 (0.536, 0.810) | 0.753 (0.694, 0.807) | 0.838 (0.793, 0.879) |
| Sensitivity | 0.766 (0.667, 0.857) | 0.714 (0.538, 0.875) | 0.819 (0.750, 0.882) | 0.727 (0.664, 0.790) |
| Specificity | 0.632 (0.542, 0.722) | 0.983 (0.898, 0.974) | 0.297 (0.188, 0.401) | 0.226 (0.058, 0.500) |
| PPV | 0.584 (0.484, 0.679) | 0.667 (0.487, 0.833) | 0.698 (0.624, 0.769) | 0.925 (0.879, 0.964) |
| NPV | 0.800 (0.714, 0.879) | 0.905 (0.914, 0.981) | 0.452 (0.300, 0.605) | 0.089 (0.020, 0.184) |
| AUC | 0.789 (0.724, 0.848) | 0.910 (0.831, 0.964) | 0.659 (0.573, 0.739) | 0.583 (0.467, 0.723) |
| AUPRC | 0.727 (0.628, 0.811) | 0.681 (0.496, 0.839) | 0.781 (0.696, 0.859) | 0.093 (0.047, 0.169) |

### 2.4. Statistical associations between primary and secondary outcomes

To better understand the relationships between the primary outcome (medication resistance) and the secondary outcomes (history of clinical relapse, history of escape, and medication non-compliance), we conducted a pairwise correlation analysis using the Phi coefficient^24^, which measures association between binary variables (-1 to 1, with values closer to ±1 indicating stronger associations). The analysis revealed a moderate positive association between medication resistance and medication non-compliance (φ = 0.213, FDR-adjusted p < 0.0001), with weaker but significant associations between medication resistance and history of clinical relapse (φ = 0.129, FDR-adjusted p < 0.0003), or history of escape (φ = 0.0930, FDR-adjusted p = 0.008).

To further investigate the strength and significance of these associations, we conducted a stratified analysis by gender and age group using Fisher’s Exact Test^25^, which does not rely on large-sample approximations. Within the male subgroup, which represented 85.2% of the sample, the associations between medication resistance and medication non-compliance (φ = 0.210, FDR-adjusted p <0.0001), history of clinical relapse (φ = 0.140, FDR-adjusted p = 0.0003), and history of escape (φ = 0.103, FDR-adjusted p = 0.006) remained statistically significant. The age-stratified analysis revealed that the associations between medication resistance and the secondary outcomes were most pronounced in the 25-55 age group, with significant associations observed for medication non-compliance (FDR-adjusted p <0.0001), history of clinical relapse (FDR-adjusted p <0.0001), and history of escape (FDR-adjusted p = 0.0288). In contrast, the associations were weaker and not statistically significant in the 16-24 and 45-76 age groups, possibly due to smaller sample sizes in these subgroups.

### 2.5. Overall feature importance patterns in psychotic disorder classifiers

To gain further insights into factors underlying model performance, we examined feature importance across all outcomes (medication resistance, clinical relapse, medication noncompliance, and escape behaviors) using the full set of 95 candidate features. By applying Random forest classifiers with impurity-corrected importance measures - an approach that helps mitigate biases favoring continuous or multi-category variables - we identified distinct importance profiles associated with each outcome (**Figure 2**). For instance, "PrevUnresponsive" (a measure of previous non-response to treatment) ranked highest for medication resistance and maintained strong influence for noncompliance (1st) and clinical relapse (5th), though its importance diminished for escape behaviors (8th). Another feature, "CurrentClinRisk," demonstrated substantial variability, ranking 2nd for medication resistance and 3rd for noncompliance but showing markedly reduced importance for escape (28th) and clinical relapse (93rd). Notably, "NumPrevAdmissions" exhibited strong importance across multiple outcomes—5th for medication resistance, 1st for clinical relapse, and 1st for escape—yet proved less critical (37th) for noncompliance. These results suggest that certain features, like "PrevUnresponsive" and "NumPrevAdmissions," exert broad influence across multiple domains, while others demonstrate outcome-specific relevance.

#### 2.5.1. Feature rankings vary between full-feature and feature transfer classifiers

Next, we evaluated how the top 30% of medication resistance features performed when transferred to predict other outcomes, comparing their importance rankings to those derived from models trained independently for each outcome (**Figure 3**). In the Feature Transfer models, certain features retained high importance across multiple outcomes. For example, "PrevUnresponsive" consistently emerged as a key predictor, ranking 1st for both medication resistance and noncompliance, 2nd for clinical relapse, and 7th for escape. Similarly, "IncapableConsentTX" remained influential across different tasks, though its relative rank varied. On the other hand, "APType2" (antipsychotic medication type), which ranked 3rd in medication resistance within the Feature Transfer framework, showed lower importance for clinical relapse (23rd) and different intermediate rankings for the other outcomes. Comparing these results with those from the full-feature Outcome-Specific models (**Figure 2**) reveals that restricting the model to the core medication resistance features produces a different hierarchy of predictors. While some features maintain their status as broadly relevant indicators, others gain or lose importance depending on the predictive context. This variability underscores the complexity of clinical risk factors and suggests that a carefully chosen subset of features can still capture essential risk dimensions, although the nature and strength of these relationships may differ depending on the specific outcome of interest.

**Figure 3.**
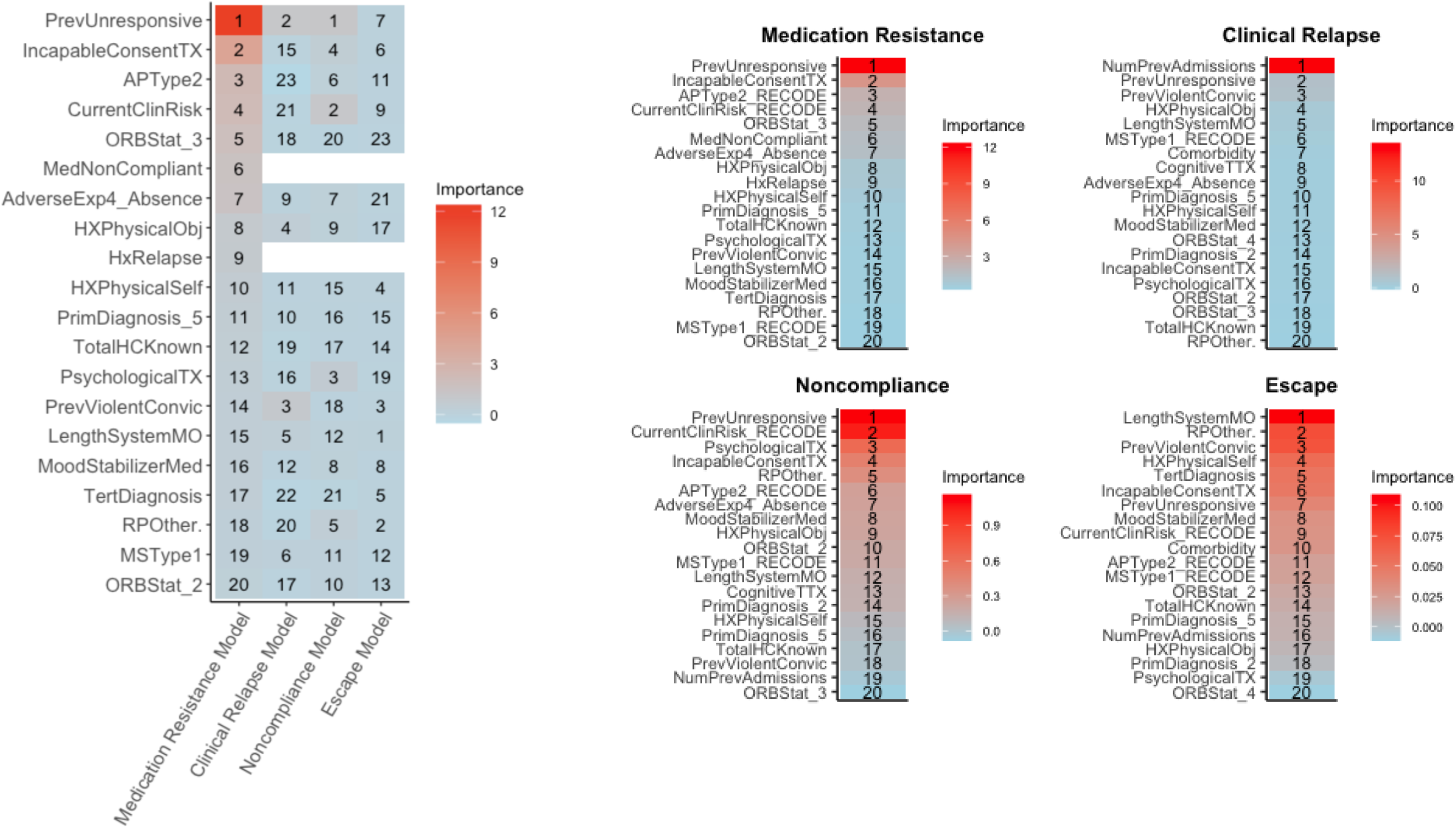
| Variable importance ranking across feature transfer models in psychotic disorders. Analysis of feature importance across four outcomes (medication resistance, clinical relapse, medication noncompliance, and escape behaviors) using the top 25 features identified through recursive feature elimination for medication resistance prediction, contrasting with the full 95-feature analysis in Figure 2. The feature set includes prior treatment response ("PrevUnresponsive"), capacity to consent to treatment ("IncapableConsentTX"), antipsychotic prescription patterns ("APType2", frequency encoded), public safety risk assessment ("CurrentClinicalRisk"), forensic status ("ORBStat_3", not criminally responsible), treatment adherence ("MedicationNoncompliant"), parental absence during development ("AdverseExperience4"), aggressive behaviors (against objects - "HxPhysicalObject", including property damage and fire-setting; against self - "HxPhysicalSelf", including self-harm and suicide attempts), diagnostic information (bipolar disorder - "PrimDiagnosis_5", psychiatric comorbidities - "TertDiagnosis"), early life instability ("TotalHXKnown", childhood home configurations), current psychological interventions ("PsychologicalTX"), criminal history ("PrevViolentConvic"), time under forensic supervision ("LengthSystemMO"), and medication profiles ("MoodStabilizerMed", "MSType1" - frequency encoded). Left panel: heatmap showing rankings of these 25 features across all outcomes, with numbers indicating specific feature rankings and warmer colors representing higher importance. Right panel: four separate heatmaps displaying importance rankings for each outcome model. Secondary outcome variables were removed from the feature set to prevent circular prediction. Among the transferred features, PrevUnresponsive" maintains high importance across outcomes, ranking first for medication resistance, second for clinical relapse, first for noncompliance, and seventh for escape behaviors. Furthermore, "IncapableConsentTX" shows consistent importance across outcomes (ranking second, twenty-third, sixth, and eleventh respectively), while "APType2" demonstrates outcome-specific importance, ranking third for medication resistance but lower for other outcomes (twenty-third, sixth, and eleventh respectively). This differential pattern of feature importance across outcomes, even within the restricted feature set, suggests that while some predictors maintain broad relevance across clinical outcomes, others may be specific to particular aspects of psychotic disorders.

#### 2.5.2. Variable importance analysis across primary and secondary outcomes in schizophrenia

In the schizophrenia subgroup (*n* = 634), variable importance analysis revealed distinct predictors for medication resistance, clinical relapse, and escape behaviors (Figure 4). Medication resistance prediction was primarily driven by treatment history (e.g., "PrevUnresponsive," ranking 1st) and clinical risk assessments ("CurrentClinRisk," 2nd). In contrast, admission history ("NumPrevAdmissions," 1st) and early illness onset markers ("AgeFirstAdmission," 3rd) drove clinical relapse prediction, while behavioral aggression ("HXPhysicalSelf," 1st) and psychiatric complexity ("TertDiagnosis," 2nd) defined escape behavior models. Compared to the broader psychotic disorders cohort (Figure 3), schizophrenia models prioritized treatment-specific predictors (e.g., antipsychotic type ["APType2"] ranked 4th vs. 19th) and aggression-related features (e.g., "HXPhysicalObj" ranked 4th vs. absent in top 20). Escape behavior prediction diverged most starkly: the broader cohort emphasized institutional history (e.g., "LengthSystemMO"), whereas schizophrenia models highlighted self-directed violence and criminal justice involvement ("PrevIncarceration").

**Figure 4.**
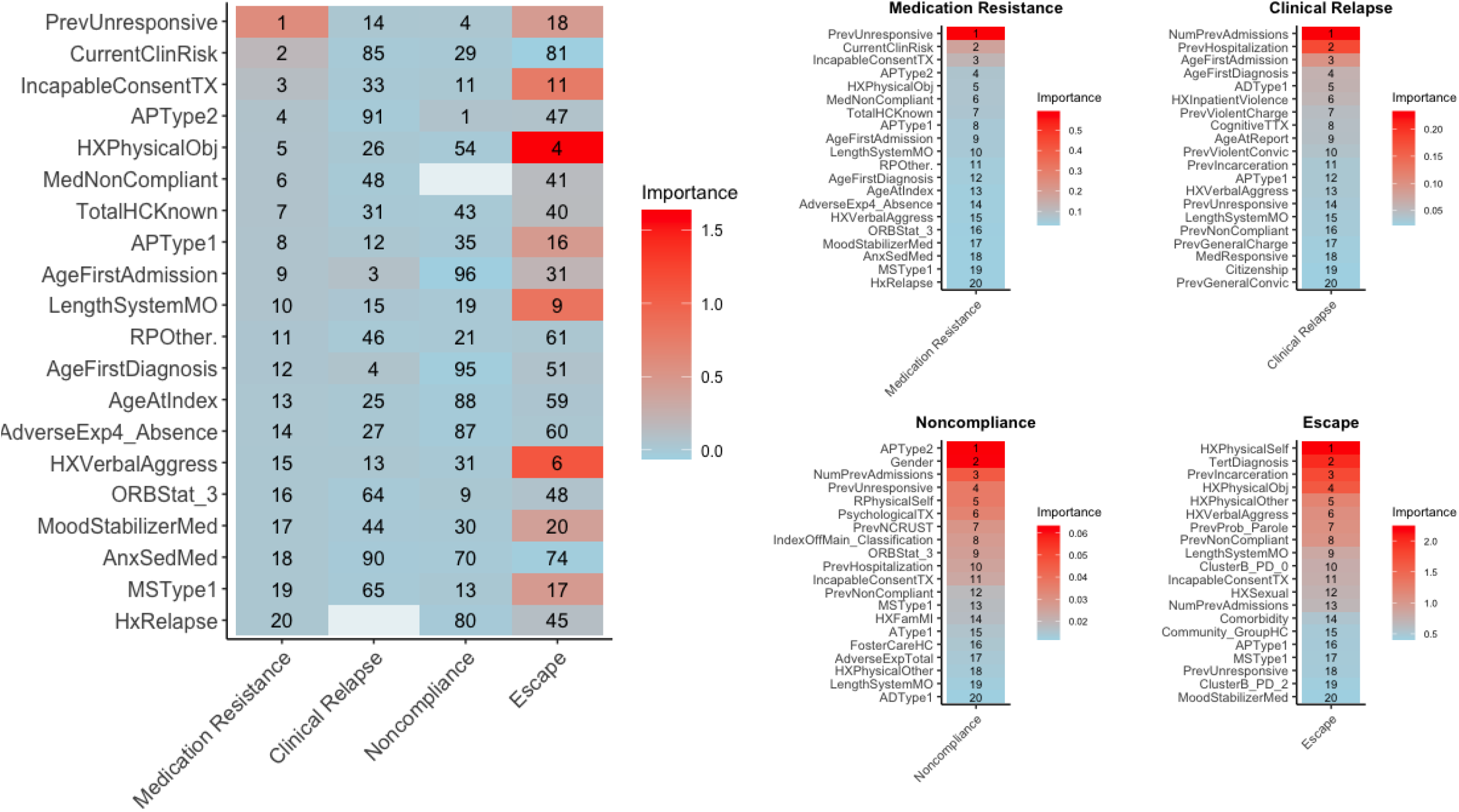
| Random forest variable importance ranking across data-driven models in Schizophrenia. Analysis of feature importance across four outcomes (medication resistance, clinical relapse, medication noncompliance, and escape behaviors) in 634 patients with schizophrenia using random forest classifiers with 95 candidate features. **Left panel**: heatmap showing the top 20 features from the medication resistance model and their rankings across outcomes, with white spaces indicating where features served as outcome variables. The features represent key clinical and demographic indicators: APType1 and APType2 indicate primary and secondary antipsychotic prescription patterns (frequency-encoded); ADType1 denotes antidepressant medication type; MSType1 represents mood stabilizer medication type; PrevUnresponsive indicates previous unresponsiveness to medication; CurrentClinRisk reflects current clinical risk assessment; NumPrevAdmissions counts previous psychiatric admissions; PrevHospitalization denotes previous psychiatric institutionalization; AgeFirstDx and AgeFirstAdmission capture age at initial diagnosis and first admission; HxInpatientViolence indicates history of violent behavior during institutionalization; PrevViolentCharge and PrevViolentConvic represent violent criminal history; CognitiveTTX denotes current cognitive remediation treatment; PrevNCRUST indicates prior NCR/UST status; HXFamMI represents family history of mental illness; PsychTx denotes current psychological treatment; TertDiagnosis indicates presence of multiple psychiatric comorbidities; ClusterB_PD reflects presence or absence of cluster B personality disorders; and Community_GroupHC indicates early life placement in community group homes. **Right panel:** Four separate heatmaps display top 20 features for each outcome model, with warmer colors indicating higher importance. In clinical relapse prediction, institutional metrics dominate, with number of previous psychiatric admissions (NumPrevAdmissions) and previous psychiatric hospitalization history (PrevHospitalization) leading the rankings. Early clinical indicators follow, including age at first psychiatric admission (AgeFirstAdmission) and first diagnosis (AgeFirstDiagnosis). Treatment response emerges through antidepressant medication type (ADType1), while historical inpatient violence patterns (HxInpatientViolence) demonstrate significant predictive value. The noncompliance model prioritizes medication profiles, with secondary antipsychotic medication type (APType2) as the primary predictor, followed by demographic factors (Gender) and institutional history (NumPrevAdmissions). Recent clinical indicators, including previous unresponsiveness to medication (PrevUnresponsive) and current self-directed violence in inpatient settings (RPhysicalSelf), precede current psychological treatment engagement (PsychologicalTX). Legal status shows substantial importance through prior findings of Not Criminally Responsible or Unfit to Stand Trial (PrevNCRUST) and current offense type (IndexOffMain_Classification). The escape model emphasizes aggressive behaviors, beginning with history of self-directed violence including self-harm and suicide attempts (HXPhysicalSelf), psychiatric complexity (multiple comorbidities; TertDiagnosis), and previous penal institution incarceration (PrevIncarceration). Various forms of institutional aggression follow, including property destruction (HXPhysicalObj), violence toward others (HXPhysicalOther), and verbal aggression (HXVerbalAggress). Criminal history through previous probation or parole status (PrevProb_Parole) and treatment adherence (PrevNonCompliant) round out the top predictors. Prior unresponsiveness to medication (PrevUnresponsive) maintains high importance across outcomes, while current clinical risk assessment (CurrentClinRisk) shows more consistent predictive value in schizophrenia compared to broader psychosis. Historical aggressive behaviors, particularly physical aggression across multiple domains, demonstrate uniquely high importance, suggesting these behavioral patterns may be especially relevant for outcome prediction in schizophrenia.

**Figure 5.**
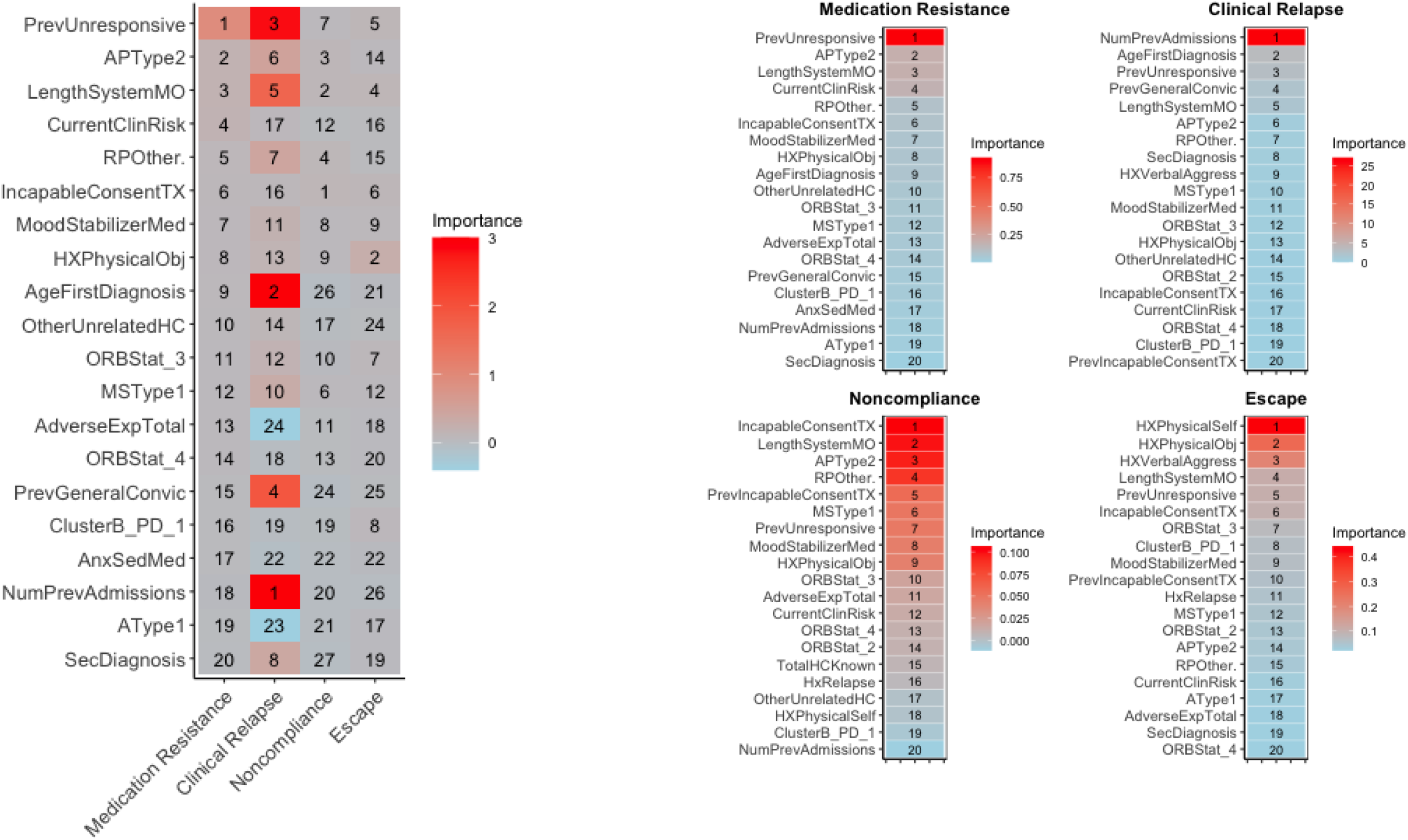
| Random forest variable importance ranking across feature transfer models in Schizophrenia. Analysis of feature importance across four clinical outcomes (medication resistance, clinical relapse, noncompliance, and escape behaviors) using the top 20 features identified through recursive feature elimination for medication resistance prediction. The feature set includes prior treatment response ("PrevUnresponsive"), capacity to consent to treatment ("IncapableConsentTX"), antipsychotic prescription patterns ("APType2", frequency encoded), public safety risk assessment ("CurrentClinicalRisk"), forensic status ("ORBStat_3", not criminally responsible), treatment adherence ("MedicationNoncompliant"), parental absence during development ("AdverseExperience4"), aggressive behaviors (against objects - "HxPhysicalObject", including property damage and fire-setting; against self - "HxPhysicalSelf", including self-harm and suicide attempts), diagnostic information (bipolar disorder - "PrimDiagnosis_5", psychiatric comorbidities - "TertDiagnosis"), early life instability ("TotalHXKnown", childhood home configurations), current psychological interventions ("PsychologicalTX"), criminal history ("PrevViolentConvic"), time under forensic supervision ("LengthSystemMO"), and medication profiles ("MoodStabilizerMed", "MSType1" - frequency encoded). Left panel: heatmap displaying the relative importance of these features across all outcomes, with numbers indicating specific feature rankings and warmer colors representing higher importance. Right panel: individual heatmaps showing importance rankings for each outcome model. Among the transferred features, "PrevUnresponsive" demonstrates strong importance across multiple outcomes, ranking first for medication resistance, third for clinical relapse, seventh for noncompliance, and fifth for escape behaviors. "NumPrevAdmissions" shows notable outcome-specific importance, ranking eighteenth for medication resistance but first for clinical relapse, while showing lower importance for noncompliance and escape. History of physical behaviors emerge as particularly important for escape prediction, with "HXPhysicalSelf" and "HXPhysicalObj" ranking first and second respectively. Treatment-related features like "APType2" and "LengthSystemMO" maintain moderate importance across outcomes, with "APType2" ranking second for medication resistance and showing varied importance across other outcomes. These patterns, when compared to the broader psychotic disorders sample (Figure 3), reveal more consistent feature importance rankings across outcomes in schizophrenia, particularly for clinical risk factors and treatment history variables. This suggests that predictive features may have more uniform utility across different clinical outcomes in schizophrenia compared to the more heterogeneous patterns observed in the broader psychotic disorders population.

#### 2.5.3. Feature rankings reveal outcome-specific predictors in schizophrenia

Feature Transfer models in schizophrenia (Figure 5) identified treatment engagement ("PrevUnresponsive," "LengthSystemMO") as cross-outcome predictors, despite their variable rankings in outcome-specific models (Figure 4). For clinical relapse, Feature Transfer prioritized medication resistance features over admission history while maintaining comparable performance (AUC = 0.912 vs. 0.925; F1 +0.119). Similarly, for treatment noncompliance, Feature Transfer elevated treatment history predictors (ranked 1st–2nd) that were deprioritized in outcome-specific models (ranked 4th and outside top 20). These shared features—absent in the broader psychotic disorders cohort (Figure 3)—suggest systemic care engagement (e.g., time in care) and treatment resistance encode illness severity trajectories unique to schizophrenia. Escape behavior prediction further validated this pattern: Feature Transfer retained behavioral aggression predictors (e.g., "HXPhysicalSelf," 1st) while incorporating treatment history features ("PrevUnresponsive," 5th) absent in outcome-specific models (ranked 18th), underscoring their broader relevance to risk stratification. This interpretation is strengthened by the quantitative performance comparisons between models, where Feature Transfer models demonstrated statistically significant improvements in F1 scores for clinical relapse prediction (difference: 0.119, 95% CI: 0.025 to 0.213) while maintaining comparable AUC values (Feature Transfer: 0.912 vs Outcome-Specific: 0.925, difference: -0.013, 95% CI: -0.089 to 0.063), indicating that medication resistance features effectively encode risk information that more extensive feature sets capture through different combinations of variables.

#### 2.5.4. Core predictive features maintain consistency across diagnostic groups despite outcome-specific variations

To contextualize these findings, comparing feature transfer patterns between schizophrenia (**Figure 5**) and the broader psychotic disorders sample (**Figure 3**) revealed some distinct characteristics in feature importance across outcomes. While both populations demonstrated successful feature transfer in clinical relapse and treatment non-compliance prediction, their importance rankings showed diagnosis-specific patterns. "PrevUnresponsive" maintained high importance in both populations, with particularly strong consistency in psychotic disorders where it ranked first for both medication resistance and noncompliance, second for clinical relapse, and seventh for escape. In schizophrenia, this feature showed a slightly different pattern, ranking first for medication resistance, third for clinical relapse, seventh for noncompliance, and fifth for escape behaviors, suggesting more uniform utility across outcomes in this population. "PrevUnresponsive" maintained high importance in both populations, with particularly strong consistency in psychotic disorders where it ranked first for both medication resistance and noncompliance, second for clinical relapse, and seventh for escape. Treatment-related features showed notable patterns, with "LengthSystemMO" maintaining relatively stable importance across outcomes in both populations. A key distinction emerged in escape prediction, where behavioral indicators ("HXPhysicalSelf" and "HXPhysicalObj") ranked as primary predictors in schizophrenia, while the psychotic disorders model relied more heavily on system engagement features. These differences suggest distinct mechanisms may underlie similar outcomes across diagnostic groups, despite the Feature Transfer approach maintaining predictive utility in both populations.

## 1. 3. Discussion

### 3.1 Feature transfer shows variable performance across clinical outcomes and diagnostic groups

We developed and evaluated a Feature Transfer methodology that leverages features initially identified as predictive of medication resistance to predict additional clinical outcomes in psychotic disorders. In a sample of 893 patients with psychotic disorders across 11 forensic psychiatric institutions, Feature Transfer models using 25 medication resistance-derived features achieved comparable performance to full-feature models (95 features) in predicting clinical relapse (F1 score difference: -0.007, 95% CI: -0.083 to 0.069). For medication noncompliance, the overlapping confidence intervals for both F1 score (difference: 0.002, 95% CI: -0.013 to 0.013) and AUC (difference: -0.022, 95% CI: -0.194 to 0.137) indicated no significant performance differences between Feature Transfer and full-feature models. Furthermore, Feature Transfer showed reduced effectiveness for escape behaviors (AUC difference: -0.160, 95% CI: -0.304 to -0.031) compared to outcome-specific models. Analysis of 634 patients with schizophrenia revealed enhanced utility of Feature Transfer, particularly for clinical relapse prediction where it demonstrated improved performance over full-feature models (F1 score difference: 0.119, 95% CI: 0.025 to 0.213) while maintaining comparable discriminative ability (Feature Transfer AUC: 0.912 vs Outcome-Specific: 0.925). These findings indicate that predictive features for medication resistance capture underlying risk dimensions that manifest differently across diagnostic groups and clinical outcomes, with particular relevance for relapse prediction in schizophrenia.

### 3.2. Feature transfer operates through direct feature selection and importance rankings

Feature Transfer operates at the feature selection level, representing a distinct approach to leveraging information across related clinical prediction tasks that differs fundamentally from traditional machine learning paradigms. By directly utilizing feature importance rankings rather than complex transformations or mappings, this methodology maintains interpretability while enabling flexible application to clinical outcomes. This emphasis on interpretable feature selection distinguishes Feature Transfer from traditional machine learning approaches to knowledge transfer. For instance, conventional transfer learning typically involves transferring model parameters or learned representations between tasks ^26^, while multi-task learning focuses on learning shared representations across simultaneously trained tasks ^27^. Although Feature Transfer shares some conceptual similarity with feature-based transfer learning methods, such as feature-representation transfer and feature-transformation transfer^28^, which aim to identify and map feature spaces between domains, its direct use of importance rankings rather than feature space mapping makes it both more interpretable and flexible in its application. This distinction is crucial for clinical applications, where maintaining the interpretability of original features is essential for clinical decision-making.

Our empirical validation through bootstrap analyses and McNemar’s tests provides insight into when and why Feature Transfer proves effective. The bootstrap results comparing Feature Transfer and Outcome-Specific models revealed that for clinical relapse in schizophrenia, Feature Transfer achieved superior F1 scores (difference: 0.119, 95% CI: 0.025 to 0.213) while maintaining comparable AUC values. This suggests that features predictive of medication resistance capture fundamental aspects of illness trajectory that manifest across multiple outcomes. However, the significant McNemar’s test results for both History of Relapse (χ2 = 12.04, p = 0.0005) and History of Escape (χ2 = 16.00, p < 0.0001) indicate that Feature Transfer and Outcome-Specific models make distinct classification decisions at the individual level, highlighting the importance of task similarity in determining transfer effectiveness. The model-agnostic nature of Feature Transfer distinguishes it from existing approaches that are tied to specific architectural choices or learning algorithms. While we implemented Feature Transfer using random forests, the approach could be readily adapted to other algorithms, including support vector machines ^29^, or simpler linear models, such as elastic net ^29,30^. This flexibility is particularly valuable in clinical settings, where different institutions or contexts might require different modeling approaches based on data availability, computational resources, or interpretability requirements. The effectiveness of Feature Transfer appears to depend on the relationship between the source task (medication resistance) and target tasks (secondary outcomes), as evidenced by its variable performance across outcomes. This pattern aligns with theoretical work in transfer learning suggesting that transfer effectiveness correlates with task similarity. However, Feature Transfer’s focus on feature importance rather than learned parameters or representations necessitates careful examination of how predictive architectures differ between full-feature and reduced-feature models, particularly when similar performance is achieved through different combinations of predictors.

### 3.3. Medication resistance features reveal shared predictive patterns and differential utility for risk prediction and monitoring

The comparison of outcome-specific and feature transfer models demonstrates that distinct predictive feature combinations can achieve equivalent performance across clinical outcomes, while revealing important implications for risk monitoring in psychotic disorders. In the broader psychotic disorders sample (Figures 2 and 3), outcome-specific models developed using the full feature set showed that different clinical outcomes relied on distinct combinations of predictors. For example, in outcome-specific models of escape behavior prediction (Figure 2), institutional history metrics such as “NumPrevAdmissions” exhibited the highest predictive importance, whereas previous treatment response (“PrevUnresponsive”) ranked eighth. In contrast, when feature transfer models were restricted to medication resistance features, they maintained similar predictive performance but adopted different architectures: “PrevUnresponsive” rose to seventh in importance and treatment-related features like “IncapableConsentTX” and “APType2” displayed heightened relative importance (Figure 3). This divergence in feature importance patterns was particularly evident in schizophrenia. Outcome-specific models (Figure 4) identified behavioral indicators—“HXPhysicalSelf,” “TertDiagnosis,” and “PrevIncarceration”—as the strongest predictors for escape behaviors, whereas feature transfer models (Figure 5) achieved comparable performance by prioritizing treatment history indicators, with “PrevUnresponsive” and “LengthSystemMO” ranking fifth and fourth, respectively, compared to their lower rankings (18th and 9th) in the outcome-specific models.

The significance of these distinct predictive patterns extends beyond mere ranking differences. In schizophrenia, the robust performance of feature transfer models in predicting clinical relapse—despite emphasizing different predictors than outcome-specific models—suggests that medication resistance features capture underlying vulnerability factors not immediately apparent through conventional risk assessment. This is particularly noteworthy given the moderate statistical associations observed between medication resistance and secondary outcomes in correlation analyses. These findings indicate that equivalent predictive performance can emerge from distinct feature combinations, challenging traditional approaches that assume unique predictor sets for different outcomes and suggesting that risk factors in psychotic disorders are more interconnected than previously recognized. In this context, medication resistance features may serve as proxies for broader illness vulnerability.

Our results also reveal important implications for medication management and risk monitoring in forensic psychiatric settings. The comparable performance of feature transfer models using medication resistance features to predict secondary outcomes indicates fundamental connections between treatment response and clinical trajectory. This is especially evident in the prediction of clinical relapse, where feature transfer models achieved performance equivalent to outcome-specific models in the psychotic disorders sample (F1 score difference: –0.007, 95% CI: –0.083 to 0.069) and even superior performance in schizophrenia (F1 score difference: 0.119, 95% CI: 0.025 to 0.213). In schizophrenia, the feature transfer approach demonstrated enhanced sensitivity in relapse prediction (0.912 vs. 0.802) while maintaining a high positive predictive value (0.969), implying that early indicators of medication resistance may serve as crucial warning signs for subsequent clinical deterioration. This improved detection, achieved solely with medication resistance-related features, suggests that closer monitoring of treatment response patterns may enable earlier identification of patients at heightened risk for clinical relapse.

However, the differential performance patterns across outcomes provide further insight into the complexity of risk dimensions. While medication resistance features effectively predicted both relapse and noncompliance, their limited utility in predicting escape behaviors—evidenced by a significantly lower AUC in psychotic disorders (–0.160, 95% CI: –0.304 to –0.031)—indicates that behavioral risks may operate through partially independent mechanisms. The divergent classification patterns revealed by McNemar’s tests, particularly for relapse prediction (χ² = 23.18, p < 0.0001 in schizophrenia), further suggest that feature transfer and outcome-specific models identify different subsets of high-risk patients. This has practical implications for risk stratification; combining predictive probabilities from both modeling approaches could provide complementary perspectives on patient risk and enable more nuanced monitoring strategies. Ultimately, these findings support an integrated approach to treatment planning wherein medication management—grounded in the predictive power of medication resistance features—serves as a cornerstone of risk prevention, while being complemented by outcome-specific monitoring strategies, particularly for risks such as escape behaviors that are less strongly associated with treatment response patterns. Regular assessment of medication resistance indicators, with adjustments to monitoring intensity and intervention thresholds based on feature transfer predictive probabilities, may therefore optimize risk management in these complex clinical populations.

### 3.4. Medication resistance features align with known treatment response patterns in schizophrenia

Our analyses identified that features selected for classifying medication resistance status showed associations with historical relapse and noncompliance in psychotic disorders, including schizophrenia. Previous unresponsiveness to treatment ("PrevUnresponsive") emerged as a predictive feature for current medication resistance classification as well as historical relapse and noncompliance. This finding aligns with longitudinal evidence that patients meeting treatment resistance criteria often demonstrate limited symptomatic improvement from initial antipsychotic therapy, with non-response patterns evident during early relapses^31–32^. These results also align with guidelines and systematic reviews identifying persistent non-response to adequate pharmacological treatment as a characteristic of treatment-resistant psychosis^35–36^.

Beyond pharmacological factors, additional features in our models corresponded with established predictors of poor clinical trajectories. The presence of childhood adversity ("AdverseExperience4") and aggression-related variables ("HxPhysicalObject," "HxPhysicalSelf") align with research linking early-life stressors and aggressive behavior to more complex illness courses^37–38^. Although substance use variables were not among our top features, previous studies have demonstrated that substance misuse can exacerbate treatment challenges, contribute to violence risk, and worsen long-term psychiatric outcomes^39–40^. The absence of these factors from our primary feature set does not rule out their importance; it may indicate that medication resistance features capture a subset of underlying vulnerabilities that interact with, but do not fully encompass, all relevant risk domains. Institutional and forensic characteristics also proved influential. Institutional and forensic characteristics, including "LengthSystemMO," "ORBStat_3," and "PrevViolentConvic," demonstrated predictive importance, consistent with evidence that forensic patients present with severe psychopathology, elevated non-adherence rates, and complex clinical-legal pathways^34,41–42^.

In our schizophrenia subgroup analysis, Feature Transfer models using medication resistance-derived features demonstrated improved classification of historical relapse compared to full-feature models. This finding is consistent with results from Demjaha et al. (2017) ^43^, who reported that a sizable fraction (23%) of first-episode psychosis patients met criteria for treatment resistance and that most of these individuals were resistant from the initiation of treatment. Their analyses identified schizophrenia diagnosis, earlier illness onset, and prominent negative symptoms as predictors of early resistance. In our analyses, "AgeFirstDiagnosis" emerged as a key predictor for medication resistance specifically in schizophrenia but not in the broader psychotic disorders sample. This diagnostic-specific finding suggests that earlier onset in schizophrenia may indicate a distinct illness pattern detectable near treatment initiation, supporting previous evidence linking age of onset to treatment response trajectories^43,44–45^.

Our analyses also revealed differential utility of medication resistance features across outcomes. While these features showed strong associations with historical relapse, their performance in classifying medication noncompliance was limited. This pattern highlights a key distinction between pharmacological resistance and what previous literature terms "pseudo-resistance" - apparent non-response arising from suboptimal adherence, insufficient dosing, or diagnostic uncertainty rather than biological refractoriness^46–47^. The limited classification accuracy for noncompliance suggests that medication resistance features may capture intrinsic non-response patterns but not behavioral factors affecting treatment adherence. This finding aligns with evidence that noncompliance and true treatment resistance reflect distinct pathophysiological processes^47^.

These results indicate several clinical implications. The identification of medication resistance features that associate with historical relapse could inform monitoring strategies, particularly in schizophrenia where diagnostic-specific patterns emerged. However, the distinction between true resistance and pseudo-resistance suggests the need for complementary approaches to address behavioral aspects of adherence. Future research should examine whether these cross-sectional associations persist longitudinally and how they might guide intervention strategies across different clinical settings. This work particularly emphasizes the importance of considering both pharmacological and behavioral factors when evaluating treatment response in psychotic disorders.

### 3.5. Methodological, population, and feature selection limitations

Several important limitations warrant consideration when interpreting these findings. First, the retrospective nature of our data introduces potential biases in outcome assessment and feature measurement. While our sample of 893 patients across 11 forensic psychiatric institutions provides high statistical power and institutional diversity, the reliance on historical records means that subtle clinical changes or undocumented factors might influence our prediction targets. The use of frequency encoding for medication classes, while capturing population-level prescription patterns and reducing feature dimensionality, may not fully represent individual treatment intensities or complex medication interactions that could affect resistance patterns. Another key methodological limitation stems from our use of historical indicators as proxy measures for clinical relapse and escape behavior outcomes. While history of clinical relapse and escape behaviors likely correlate with future risk, this approach assumes temporal stability in risk factors that may not hold across different stages of illness or treatment. This limitation particularly affects our ability to establish causal relationships between medication resistance and subsequent outcomes, as historical indicators cannot definitively establish temporal precedence.

The focus on forensic psychiatric patients introduces important considerations for interpretation and generalizability. This population showed substantially higher rates of treatment resistance (42.3%) compared to general psychiatric settings (20-30%)^47^, along with distinct characteristics including higher rates of previous incarceration (63.4%) and physical aggression (58.7%). Forensic patients have been shown to present with more severe symptomatology across all four Positive and Negative Syndrome Scale (PANSS) subscales^3^ and present with higher rates of treatment non-adherence relative to non-incarcerated individuals^48^. However, a large-scale Finnish study of 29,956 individuals with schizophrenia found that primary medication non-adherence is also common in the general psychiatric population, with 31.7% of patients not dispensing prescribed antipsychotics within one year^49^. Together, these findings underscore the widespread challenges of medication adherence across psychiatric populations, while highlighting the particularly complex clinical needs of forensic psychiatric patients.

Additionally, our feature selection process, while systematic, presents with some limitations. The use of recursive feature elimination with random forests may not fully capture complex non-linear interactions between features, particularly given our sample size relative to the number of potential feature combinations. The selection of 25 features, while improving model interpretability, was somewhat arbitrary and alternative thresholds might yield different transfer patterns. Additionally, the imputation of missing values using MissForest^50^, while appropriate for mixed-type data, may introduce subtle biases in feature importance rankings. Furthermore, the significant classification differences revealed by McNemar’s tests suggest our models may be sensitive to specific subgroups within the patient population, particularly evident in the stronger performance for schizophrenia compared to broader psychotic disorders. This diagnostic heterogeneity effect, combined with our focus on medication resistance as the primary outcome, might overlook alternative feature combinations that could better predict specific secondary outcomes. Additionally, while our data preprocessing excluded medication dosage information due to high variability, this omission potentially limits our ability to detect dose-dependent aspects of medication resistance. The categorical treatment of diagnostic variables, while necessary for analysis, may oversimplify complex diagnostic presentations that could influence treatment response patterns.

### 3.6. Clinical integration and healthcare system implications

Our findings demonstrate that features originally identified as predictive of medication resistance carry prognostic value for additional clinical outcomes—such as clinical relapse and medication noncompliance—in psychotic disorders. This suggests that routinely tracking key indicators (e.g., medication adherence, symptom fluctuations, and adverse events) can form the basis of dynamic risk assessment frameworks. For clinical integration, effective use of Feature Transfer-based models will require structured procedures for periodic updating of predictor variables and continuous monitoring of patient response trajectories, ensuring that decision thresholds remain current in rapidly evolving clinical settings.

Although the utility of Feature Transfer was most pronounced for schizophrenia, this diagnostic specificity underscores the need for a carefully phased implementation strategy, beginning with the subgroups that yield the most robust predictive patterns. Finally, given that Feature Transfer and outcome-specific models may identify distinct high-risk profiles, integrating both approaches may capture a more complete picture of patient risk and thus optimize resource allocation and treatment planning. Finally, longitudinal studies are needed to evaluate how these predictive patterns evolve over time and to validate the models against additional endpoints (e.g., functional recovery and symptom progression) across diverse healthcare settings. Such studies could also assess the stability of these feature patterns under different treatment protocols and documentation standards, ultimately informing clinical practice and policy in forensic psychiatric settings.

In conclusion, our study demonstrates that a Feature Transfer approach—leveraging a reduced set of medication resistance–derived predictors—achieves classification performance comparable to full-feature models across multiple clinical outcomes in psychotic disorders. Notably, in our forensic psychiatric sample and particularly within the schizophrenia subgroup, core treatment response features emerged as strong predictors of clinical relapse and noncompliance, whereas their relatively weaker performance for escape behaviors implies that distinct mechanisms underlie different risk domains. These results underscore that medication resistance features capture broader risk dimensions than conventionally assumed, and that cross-task generalization via rank-aggregated feature selection not only preserves interpretability but also provides novel insights into shared risk mechanisms. While our findings offer a promising foundation for refining risk stratification, the retrospective nature of our data and the unique characteristics of our study population necessitate prospective, multicenter, longitudinal investigations to validate these predictive patterns and elucidate their clinical impact. Ultimately, this work lays the groundwork for integrating transparent, model-agnostic predictive tools into clinical decision support systems, thereby advancing personalized intervention strategies in complex psychiatric populations.

## 1. 4. Methods

### 4.1. Data preprocessing

This study analyzed data from the Ontario Review Board (ORB)^51^, encompassing all patients with psychotic disorders deemed not criminally responsible or unfit to stand trial between 2015-2016 across 11 forensic psychiatric institutions in Ontario, Canada. The total sample comprised 893 patients with psychotic disorders, including 634 patients diagnosed with schizophrenia. First, we removed variables containing identifiable information (e.g., subject codes, dates, institution names) and medication dosage data due to high variability both within and across medication classes. Instead, we retained categorical indicators of medication classes (antipsychotics, mood stabilizers, antidepressants) to capture treatment profiles. These medication classes and diagnostic variables underwent frequency encoding, where categories were replaced with their relative frequency in the dataset. This encoding was performed separately on training and testing sets to maintain data independence while preserving information about treatment and diagnostic patterns. The final feature set (*n* = 95) incorporated these encoded variables alongside sociodemographic characteristics, clinical history, violence risk factors, psychiatric comorbidities, and psychosis-specific clinical indicators. Further details can be found in Supplementary Table S1.

To prevent circularity in outcome prediction, we implemented outcome-specific feature exclusions. While the medication resistance model utilized all features (*n* = 95), models for secondary outcomes (clinical relapse, medication non-compliance, escape behaviors) excluded both the medication responsiveness indicator and other secondary outcomes from their respective feature sets. This exclusion prevents artificial performance inflation that could arise from direct outcome relationships. For instance, medication non-compliance may influence both medication resistance and clinical relapse, necessitating its exclusion when predicting these outcomes to ensure model predictions reflect underlying patient characteristics rather than outcome correlations. Data partitioning employed stratified sampling (70% training, 30% testing) via the initial_split function within the ‘rsample’ package^52^, maintaining outcome distributions across sets. Missing value imputation utilized the MissForest algorithm^50^, selected for its ability to handle mixed-type data while preserving complex variable relationships. To prevent data leakage during imputation, we first imputed the training set and then used it as a reference for test set imputation, ensuring consistent handling of missing values while maintaining set independence.

### 4.2. Outcome variables

Our analysis examined four clinical outcomes in patients with psychotic disorders within forensic settings. The primary outcome, medication resistance, was defined as absence of symptom improvement during the current reporting period despite pharmacological treatment. For secondary outcomes, we utilized historical indicators as proxies for ongoing risk, given the absence of concurrent outcome data during the reporting period. History of relapse was defined as any previous instance of readmission due to condition deterioration or breach of conditions following institutional discharge. History of escape/absconding included both successful incidents and attempts to leave institutional care without authorization or failure to return from community passes. Treatment non-compliance indicated current non-adherence to prescribed pharmacological interventions at the time of assessment. Each outcome was coded binarily: medication resistance (0=responsive, 1=resistant), history of relapse (0=no, 1=yes), history of escape/absconding (0=no, 1=yes), and treatment compliance (0=compliant, 1=non-compliant). This coding scheme maintained consistency across prediction tasks while reflecting clinically meaningful distinctions in patient status and risk.

The distribution of outcomes across the dataset revealed varying class proportions. For medication resistance, 355 patients (39.8%) demonstrated resistance (coded as 0) while 538 (60.2%) showed responsiveness (coded as 1). This distribution was maintained in both training (248/376; 39.7% resistant) and testing (107/162; 39.8% resistant) sets. History of relapse comprised 773 patients (86.6%) with relapse history and 120 (13.4%) without, with this pattern preserved in training (543/81; 87.0% with relapse) and testing (230/39; 85.5% with relapse) partitions. Treatment non-compliance included 79 patients (8.8%) classified as non-compliant and 814 (91.2%) as compliant, maintained across training (57/567; 9.1% non-compliant) and testing (22/247; 8.2% non-compliant) sets. Given the marked class imbalance in the treatment non-compliance outcome, we implemented majority class downsampling (ratio=0.3) during model training while preserving the original distribution in the test set. History of escape showed 308 patients (34.5%) with escape history versus 585 (65.5%) without, maintained across training (220/404; 35.2% with escape history) and testing (88/181; 32.7% with escape history) sets.

### 4.3. Model Building and Evaluation

#### 4.3.1. Random forest classifiers

We implemented random forest binary classifiers using the ranger package in R to discriminate medication resistance from medication responders across both the full psychotic disorders sample and the subgroup analysis of patients with schizophrenia. Feature selection employed recursive feature elimination (RFE) via the caret package, using 5-fold cross-validation with 5 repeats to identify the top 25 predictors. RFE was selected for its ability to capture interaction effects and identify features that contribute to model performance through complex relationships rather than individual predictive strength. Hyperparameter tuning was performed using Bayesian Optimization^53^ via the mlr3 ecosystem in R^54^, tuning key parameters including mtry, min.node.size, sample.fraction, splitrule, num.trees, alpha, minprop, and max.depth. The optimization process incorporated 5-fold cross-validation with a 50-iteration limit. To address outcome class imbalance, we applied training set-derived class weights during model fitting.

Performance evaluation compared models using the full feature set (*n* = 95) against those using RFE-selected features (*n* = 25). Metrics included accuracy, sensitivity, specificity, positive and negative predictive values, F1 score, and area under both ROC and precision-recall curves. We calculated 95% confidence intervals using method-appropriate tests: binomial for accuracy, proportion tests for sensitivity and specificity, and bootstrap for AUC metrics. Feature importance was assessed using ranger’s impurity-corrected importance measure ^18^ to mitigate bias toward continuous variables and multi-category factors. Secondary outcome models (medication non-compliance, clinical relapse, escape behaviors) were developed using both the complete eligible feature set (excluding other outcomes as predictors) and the Feature Transfer approach. These parallel analyses enabled direct comparison between outcome-specific models using all available features and transfer models restricted to features identified as important for medication resistance. This framework allowed us to evaluate whether medication resistance predictors could effectively generalize to related clinical outcomes while maintaining predictive performance.

#### 4.3.2. Super learner – medication resistance

To benchmark the maximum achievable predictive performance with transferred features, we implemented a super learner ensemble binary classifier^19^ using mlr3^54^. Super learners optimize prediction by combining multiple algorithms through V-fold cross-validation, theoretically performing asymptotically as well as the best possible prediction algorithm in the user-supplied library^55^. Our implementation included six base learners selected to capture different underlying data structures: random forest (non-linear interactions)^16^, Bayesian Adaptive Regression Trees (flexible non-parametric relationships) ^56^, Support Vector Machine (complex decision boundaries)^29^, Elastic Net (sparse linear relationships)^30^, k-Nearest Neighbors (local patterns)^57^, and Naive Bayes (probabilistic classification)^58^. Each base learner underwent hyperparameter optimization via Bayesian optimization using ‘mlr3tuning’ in mlr3^54^. The meta-learner, implemented as XGBoost with tuned learning rate, tree depth, and regularization parameters, combined base learner predictions.

The super learner framework used 5-fold cross-validation, where base learners generated out-of-fold predictions on held-out data to form the meta-features for the meta-learner. This approach prevents overfitting by ensuring the meta-learner only sees predictions made on observations not used in training the base learners. The final ensemble was evaluated on a stratified 30% test set to assess generalization performance. While super learners can achieve optimal prediction performance, their multi-layer architecture limits feature importance interpretation. Although methods such as SHapley Additive exPlanations (SHAP)^59^ can be used to quantify the contribution of base learners to final predictions, they cannot directly illuminate the relationship between original features and outcomes. Given these constraints, we used the super learner primarily to validate the predictive capacity of our feature sets, while relying on random forests for feature importance analysis. The super learner framework served two key purposes in evaluating Feature Transfer efficacy. First, by applying super learner to the reduced feature set identified through Feature Transfer, we established an upper bound on achievable predictive performance using only medication resistance-derived features. Second, comparing super learner and random forest performance on these features provided validation that random forest models sufficiently captured predictive relationships for feature importance interpretation. This dual application of super learner and random forest classifiers enabled assessment of both maximum predictive capacity and feature-level insights.

#### 4.3.3. Feature transfer

The Feature Transfer framework evaluates whether predictors of medication resistance generalize to three secondary outcomes: clinical relapse, treatment non-adherence, and escape behaviors. By transferring the ranked importance of predictors from the medication resistance model, this framework allows us to directly assess whether these variables retain their predictive relevance when applied to related outcomes. In doing so, it provides a transparent means to compare the impact of a streamlined, interpretable feature set against traditional, full-feature models. This approach prioritizes feature importance transfer over parameter-based transfer learning, retaining interpretability while testing the hypothesis that clinical, demographic, and environmental factors exhibit cross-outcome predictive relevance. For medication resistance prediction, recursive feature elimination (RFE; see Section 4.3.1) identified the top 25 predictors using random forest classifiers and impurity-corrected importance measures. These features—selected for their capacity to model non-linear relationships—were then used to train new random forest classifiers for secondary outcomes. Parallel outcome-specific models were developed using all 95 features to establish baseline performance. Both approaches employed identical hyperparameter optimization and cross-validation strategies (5-fold, 5 repeats).

To further validate the generalizability of the selected features, we conducted multiple analyses. First, we compared the distributions of the selected features across different outcomes using summary statistics, visual plots, and the Kolmogorov-Smirnov test^60^ to identify outcome-specific variations. Next, correlation analysis (using phi coefficients) and comparative evaluations of feature importance rankings between Feature Transfer and Outcome-Specific models were performed to examine consistency. Performance equivalence between modeling approaches was then assessed through stratified bootstrap analysis^61^ with 10,000 resamples, preserving original outcome distributions while evaluating the reduction from 95 to 25 features. For each resample, differences in AUC-ROC and F1 scores between paired models were computed to estimate 95% confidence intervals for true performance differences, where p-values were computed as the proportion of resamples in which the absolute difference exceeded the observed difference.

To complement the bootstrap analysis, case-level classification differences between Feature Transfer and Outcome-Specific models were evaluated using McNemar’s test with Edwards’ continuity correction^62^. This test examined asymmetry in classification decisions between paired models by constructing a 2×2 contingency table of discordant predictions, where entries represented cases correctly classified by one model but misclassified by the other^23^. The test statistic followed a chi-square distribution under the null hypothesis of symmetric disagreement patterns, with statistical significance assessed at α = 0.05. The Benjamini-Hochberg procedure was applied separately to three families of tests (bootstrap, McNemar’s, and Kolmogorov-Smirnov^60^) to control the false discovery rate. These analyses were carried out separately for the full sample (n=893) and a subgroup analysis of patients with schizophrenia (n=634).

## Data Availability

The data used in this study were derived from deidentified electronic health records and Ontario Review Board records accessed through St. Joseph's Healthcare Hamilton. Due to the sensitive nature of forensic psychiatric data and institutional privacy requirements, the dataset cannot be made publicly available. Qualified researchers may request access to a de-identified dataset by contacting the Research Ethics Board at St. Joseph's Healthcare Hamilton, with appropriate institutional and ethical approvals.

