## supplementary Figure 1 for "Leveraging Feature Transfer to Predict Medication Resistance and Secondary-Clinical Outcomes in Psychotic Disorders in Forensic Settings"

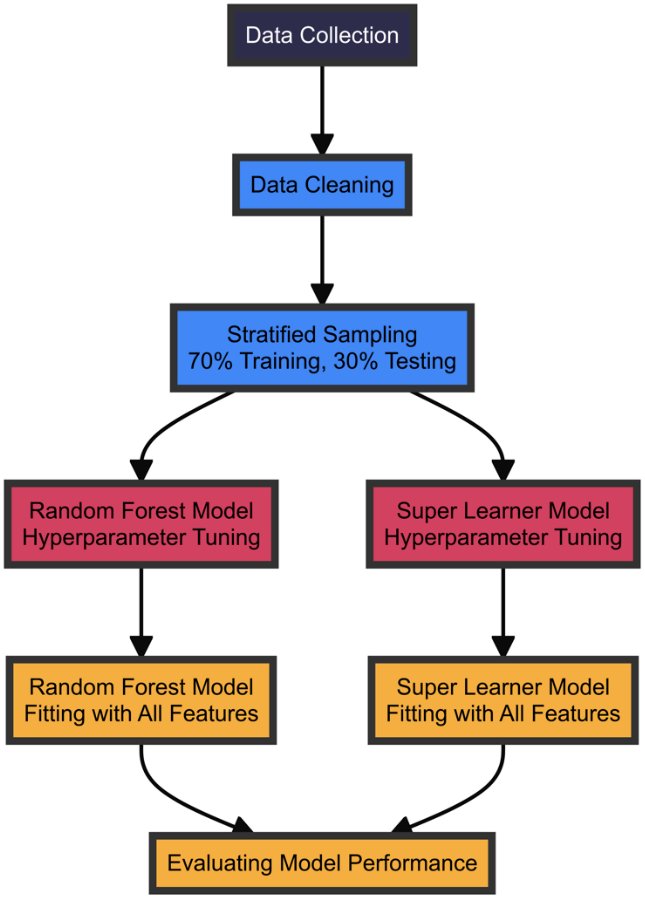


**Supplementary Figure 1 | Outcome-specific modeling framework.** An illustration of the data-driven modeling framework used to develop and evaluate our Outcome-Specific models, which serve as a baseline for comparison with the Feature Transfer models discussed in Figure 1. This framework was applied to four distinct outcomes: medication resistance (primary), history of clinical relapse, history of escape behaviors, and medication noncompliance (secondary). The process begins with data collection, followed by data cleaning to ensure quality and consistency as described in the Methods. The cleaned dataset is then divided into training (seventy percent) and testing (thirty percent) sets using stratified sampling to maintain outcome distribution. Two modeling approaches are employed: Random Forest and Super Learner. For each approach, hyperparameter tuning is performed using Bayesian Optimization, followed by model fitting using all ninety-five candidate features. By using all candidate features, the Outcome-Specific models offer a fair comparison to the Feature Transfer models. This comparison allows us to assess whether the transferred features, despite being fewer in number, maintain predictive power across different outcomes. The systematic comparison of feature importance and performance metrics between these two approaches provides insights into the generalizability and efficiency of the Feature Transfer methodology. The performance of both Random Forest and Super Learner models are then evaluated using various metrics including sensitivity, specificity, PPV, NPV, AUC and AUPRC. This approach provides a robust baseline against which to compare our Feature Transfer models and ascertain whether there are key predictors that are relevant across multiple psychiatric outcomes.
