## Supplementary figures and images for "Leveraging Feature Transfer to Predict Medication Resistance and Secondary-Clinical Outcomes in Psychotic Disorders in Forensic Settings"

### Supplementary Figure 2

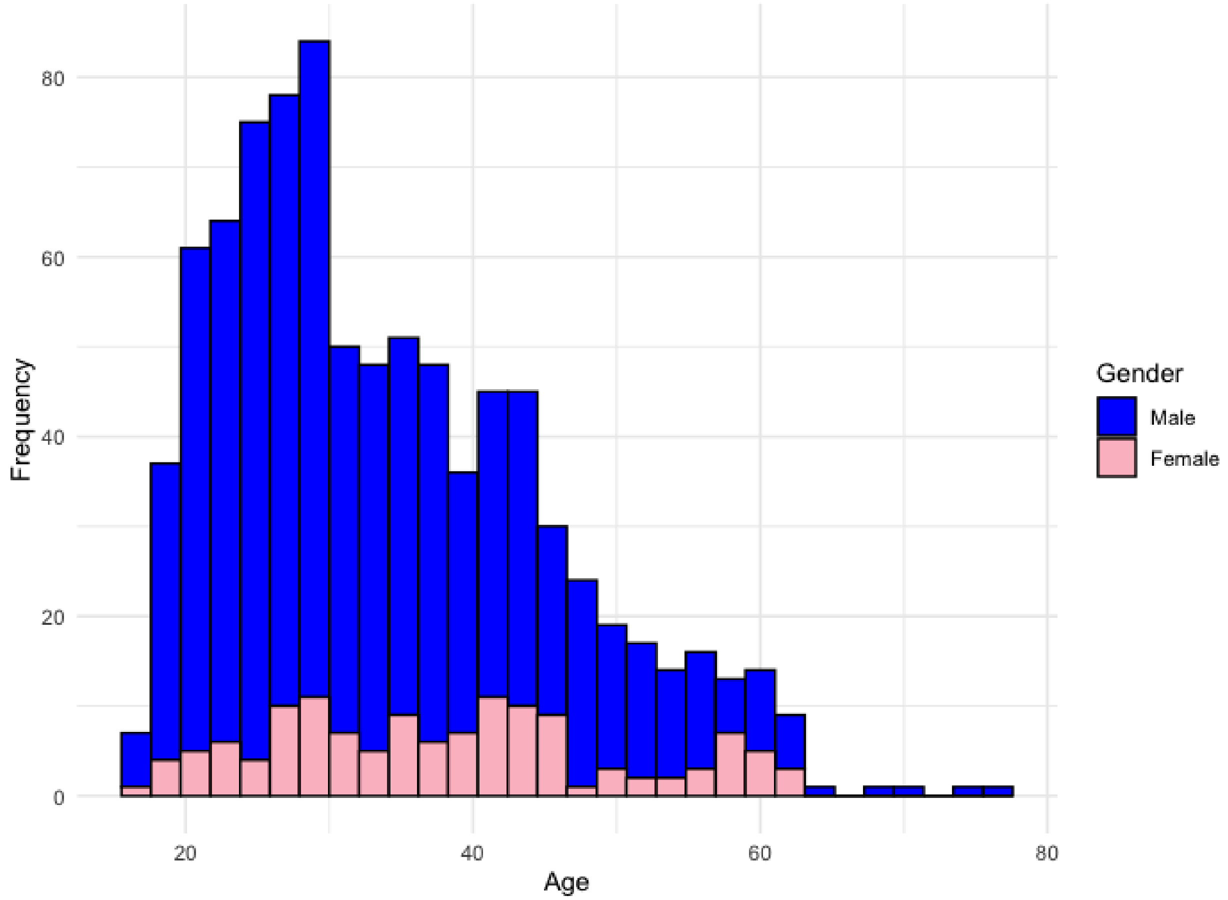


Supplementary Figure 2 | Sample age distribution stratified by gender.
